# Brain cross-omics integration in Alzheimer’s disease

**DOI:** 10.1101/2022.12.10.22283295

**Authors:** Abdallah M. Eteleeb, Brenna C. Novotny, Carolina Soriano Tarraga, Christopher Sohn, Eliza Dhungel, Logan Brase, Aasritha Nallapu, Jared Buss, Fabiana Farias, Kristy Bergmann, Joseph Bradley, Joanne Norton, Jen Gentsch, Fengxian Wang, Albert A. Davis, John C. Morris, Celeste M. Karch, Richard J. Perrin, Bruno A. Benitez, Oscar Harari

## Abstract

Unbiased data-driven omic approaches are revealing the molecular heterogeneity of Alzheimer disease. Here, we used machine learning approaches to integrate high-throughput bulk and single-nucleus transcriptomic, proteomic, metabolomic, and lipidomic profiles with clinical and neuropathological data from multiple AD cohorts. We discovered four unique multimodal molecular profiles, one showing signs of poor cognitive function, a faster pace of disease progression, shorter survival with the disease, severe neurodegeneration and astrogliosis, and reduced levels of metabolomic profiles. This profile shows similar cellular and molecular profiles in multiple affected cortical regions associated with higher Braak tau scores and significant dysregulation of synapse-related genes and endocytosis, phagosome, mTOR signaling pathways altered in AD early and late stages. The multimodal clusters uncovered cerebrospinal fluid biomarkers to monitor AD progression. AD cross-omics data integration with transcriptomic data from an SNCA mouse model revealed an overlapping signature. Our cross-omics analyses provide novel critical molecular insights into AD.

## Introduction

Alzheimer disease (AD) is a heterogeneous multifactorial neurodegenerative disorder pathologically characterized by amyloid (Aβ) plaques, neurofibrillary tangles (NFTs), neuroinflammation, and synaptic and neuronal loss. Recently, multiple clinical and pathological AD subtypes have been identified based on distinct spatiotemporal trajectories of tau pathology, brain atrophy, postmortem brain transcriptomics profiles, or cerebrospinal fluid proteomics. Differences in characteristics associated with these subtypes including age at onset, sex, APOE ℇ4 carrier status, cognitive status, disease duration, and cerebrospinal fluid (CSF) biomarker levels, were previously reported [1–9]. Related research efforts have leveraged molecular profiles and biomarkers to characterize and cluster the AD population into subgroups and to provide aspects of AD progression that, in turn, could inform clinical trials besides cognitive function or amyloid tau biomarkers. Novel biomarkers for AD biological processes and cognition, in general, can shape new clinical trials and are excruciatingly needed for those therapies targeting amyloid plaques of tau fibrils. However, the influence of such AD profiles on cognitive decline remains poorly understood and under-studied, representing a barrier to using this information effectively in clinical trials of potential disease-modifying therapies. In addition, previous studies have proposed multiple AD subtypes based on molecular features [5]. Despite the substantial effort in identifying molecular subtypes of AD described in this study, broader conclusions are limited by its nature as single-omic analyses typically capture changes in a single component of the biological cascade.

We have previously leveraged high-throughput brain molecular data from AD patients and healthy individuals using transcriptomic [10–12], proteomic [13], metabolomic [14] and single-cell omics [15] to study the signatures for AD. However, we recognize that omics layers are interdependent and interconnected. It has been hypothesized that by combining multiple signals from various molecules, novel significant biological hubs or pathway changes in AD will emerge that are otherwise missed. Previous studies that aggregate multiple modalities of omics (cross-omics) have proven to be a powerful approach to identifying molecular subtypes for complex human diseases, including cancer [16–21]. Different modalities of omics layers describe a biological system at different biomolecular levels [22], allowing more comprehensive disease segregation into molecular profiles that cannot be appreciated through an analysis restricted to a single modality. Cross-omics approaches can reveal how complex biomolecular profiles change in association with a disease and the relationships and correlations between distinct classes of biological molecules. Their potential for studying neurodegeneration has been previously described [23–32]. However, only recently has the deep molecular characterization of multiple cortical regions from diverse brain banks and cohorts been generated [2,3], enabling a more detailed and comprehensive examination of cross-omics data in AD.

In this study, we sought to investigate whether cross-omics studies of AD brains can provide novel molecular insights into the progression of AD and its staging information. We leveraged machine learning, digital deconvolution, and traditional statistical approaches to integrate and analyze multiple high-throughput *omics* datasets from different cortical regions and cohorts. Our cross-omics data integration approach identified four distinct molecular profiles of AD, one of which was associated with worse cognitive function and neuropathological features, including significantly higher Clinical Dementia Rating (CDR) at death, shorter survival after symptom onset, more severe neurodegeneration and astrogliosis, and decreased levels of metabolomic profiles. The molecular signatures of this profile, present in multiple cortical regions, show a significant dysregulation of synapse-related genes and pathways, suggesting neuron/synapse losses and dysfunction at later stages of AD. Integrating cross-omics approaches with data from established mouse models of neurodegeneration clarified the role of these genes and pathways in the AD pathophysiology. Furthermore, combining cross-omics data with single-cell resolution uncovered molecular profiles associated with AD features to an unprecedented level of granularity and complexity. Subsequent AD staging and network analyses, as well as survival analyses of CSF proteins related to the synapses, revealed dysregulated of synaptic proteins associated with an increased risk of dementia progression including *IGF1*, *NRXN3*, and *YWHAZ* that can be employed as possible biomarkers for early synaptic dysfunction, cognition, and AD staging.

Our results suggest that cross-omics analyses capture molecular heterogeneity among brains from AD cases that are otherwise missed in non-clustered single-modality *omics* approaches, and demonstrate that, under molecular stratification, cross-omics approaches provide new perspectives of molecular pathways associated with AD that are reflected in associations of cognitive decline progression and peripheral tissue. These novel molecular findings may open the possibility for new biomarkers for the molecular staging of AD, as well as potential therapeutic targets to ameliorate cognitive decline and AD progression that may provide a foundation for implementing precision medicine approaches for AD.

## Results

### Study design

We leveraged high-throughput transcriptomic (Single-nucleus and bulk RNA-seq), proteomic, metabolomic, and lipidomic data from the parietal cortex of participants with neuropathology-confirmed AD from the Knight Alzheimer Disease Research Center (Knight ADRC) for our discovery cohort (see Methods, **Fig 1** and **S1 Table**). Omics data were generated using next-generation sequencing/10X genomics, SomaScan, and Metabolon Precision Metabolomics platforms for transcriptomics (60,0754 transcripts) [10–12,15], proteomics (1,092 proteins) [13], and metabolomics (627 metabolites) [14], respectively (see Methods). Preprocessing, quality control (QC), and feature selection were performed on each omics data type, and only samples that passed the QC criteria in all three omics modalities were retained, resulting in 278 samples (255 AD and 23 control). We employed a Bayesian integrative clustering method (iClusterBayes [33]) to integrate AD multi-omics data (see Methods) and identify molecular profiles of AD. Control cases were later integrated for comparative analyses.

**Fig. 1.**
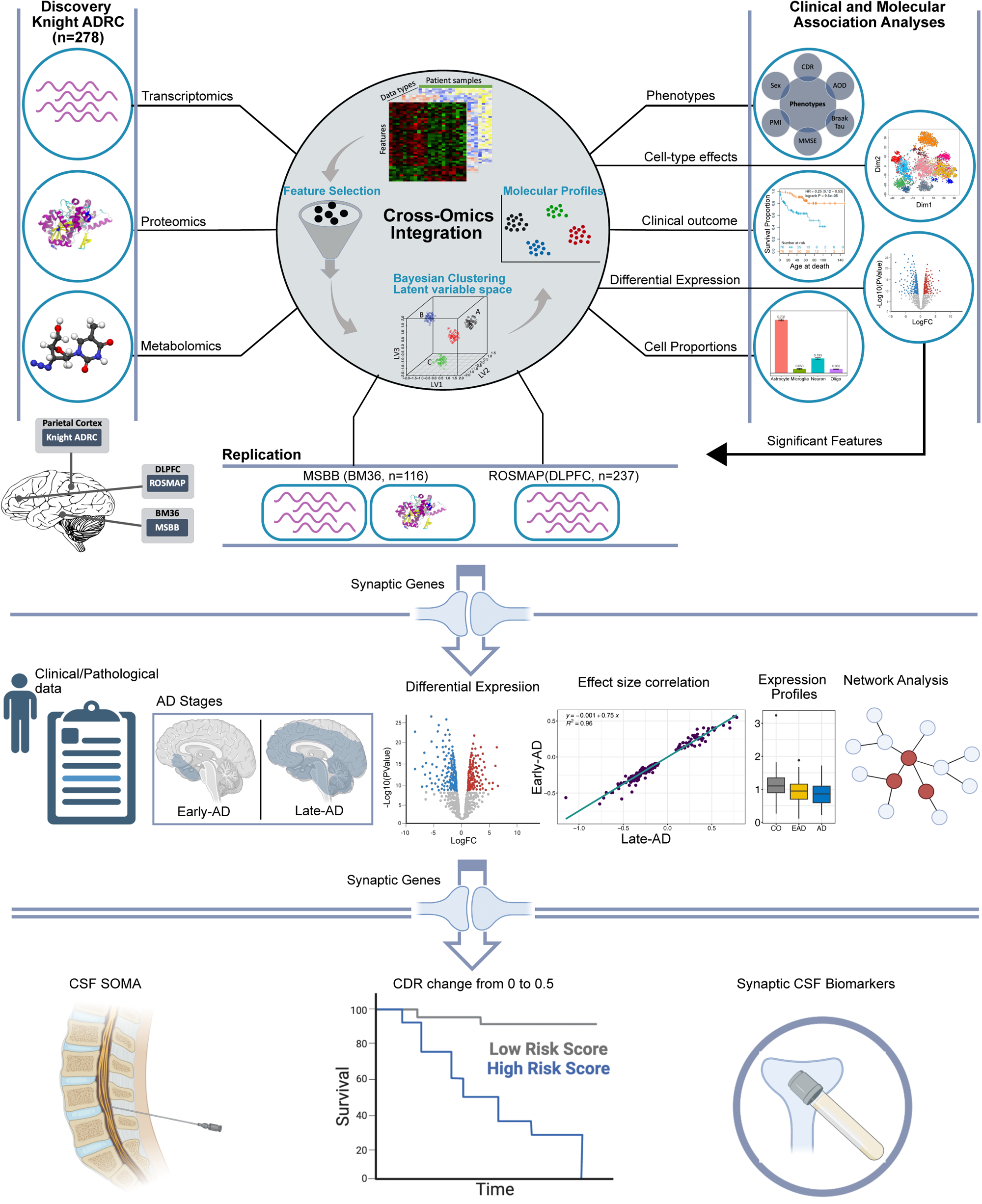
Study design. The transcriptomics, proteomics, metabolomics, and lipidomics profiles of postmortem parietal cortex samples from the Knight ADRC participants were used. The three omics shared two hundred seventy-eight subjects (255 sporadic AD and 23 control cases). Expression/reading matrices were prepared for the same set of samples (n=278) and different numbers of features (genes=60,754, proteins=1,092, metabolites=627). Before the integration, feature selection was performed on all omics datasets whose number of features exceeded 1000 features by selecting the top and most variant features. A Bayesian integrative clustering method was then employed to integrate the three omics datasets, and the best clustering solution was extracted based on the maximum deviance ratio and the minimum Bayesian information criterion (BIC). AD molecular profiles were then linked to multiple clinical and molecular attributes to examine whether an association with these attributes exists. In addition, multiple analyses, including survival, differential expression, cell-type specific effect, and cell proportions inference analyses, were performed on these profiles. These molecular profiles were then replicated in two independent datasets from MSBB BM36 (AD=93, CO=23) and ROSMAP (AD=144, CO=93). Dysregulated synaptic genes in the Knight-C4 profile were extracted and used for downstream AD staging analyses and in the ROSMAP cohort (early-AD vs. control and late-AD vs. control). Common synaptic genes present in the CSF SOMA dataset and associated with an increased risk of dementia through survival analyses were identified as CSF synaptic biomarkers for AD staging.

We next performed multiple association analyses, including association with clinical and neuropathological attributes (e.g., sex, age of death, age at onset, post-mortem interval, CDR, Braak amyloid stage, Braak neurofibrillary tangle stage), survival, and differential expression analyses to characterize AD molecular profiles. We leveraged digital deconvolution approaches to infer the cellular population structure using bulk RNA-seq data [12], and tested for association of these profiles with cell-type abundances. We also integrated single-nuclei RNA-seq data (snRNA-seq) from the parietal cortex of 67 Knight ADRC participants to determine the specific cell-type effects.

For replication, we leveraged transcriptomics (bulk RNA-seq) and proteomics (tandem mass tag; TMT) data from the parahippocampal gyri (BM36) of 116 brains (93 AD and 23 control, **S1 Table**) from The Mount Sinai Brain Bank (MSBB) study [34] cohort, as well as transcriptomics data from 237 brains (144 AD and 93 control, **S1 Table**) dorsolateral prefrontal cortex (DLPFC) samples from The Religious Orders Study and Memory and Aging Project (ROSMAP) cohort [35], accessed through the AMP-AD Synapse portal. Preprocessing, QC, and feature selection were performed like that used to generate the discovery datasets.

Furthermore, we stratified AD cases into early and late-stage AD based on clinical and neuropathological rules from ROSMAP study (see Methods, **Fig 1**) and performed DE and co-expression network analyses to identify the magnitude of the dysregulation of genes at distinct AD stages and provided a deeper understanding of the underlying dynamics of AD and approximate its progression over time (**Fig 1**). Similarly, we evaluated the association of protein levels encoded by selected genes in CSF and disease risk progression (CDR change from 0 to 0.5) using survival analyses in CSF proteomic data from the Knight ADRC participants (**Fig 1**).

### A multimodal profile of AD brains is associated with poor cognitive function and molecular attributes

We initially identified distinct AD molecular profiles by optimizing the number of clusters needed to capture the molecular heterogeneity present in AD brains from the Knight ADRC, independently of any neuropathological or clinical data. (**S1 Fig**). We identified four clusters (**Fig 2A**) in which the molecular profiles of Cluster4 (named Knight-C4 hereafter, n=42) exhibited a pronounced dysregulation of genes, proteins, and metabolites. For instance, the transcriptomic profiles of the top 75% of quantile genes that contribute significantly to the clustering solution show significant dysregulation signatures associated with Knight-C4 (**Fig 2B**).

**Fig. 2.**
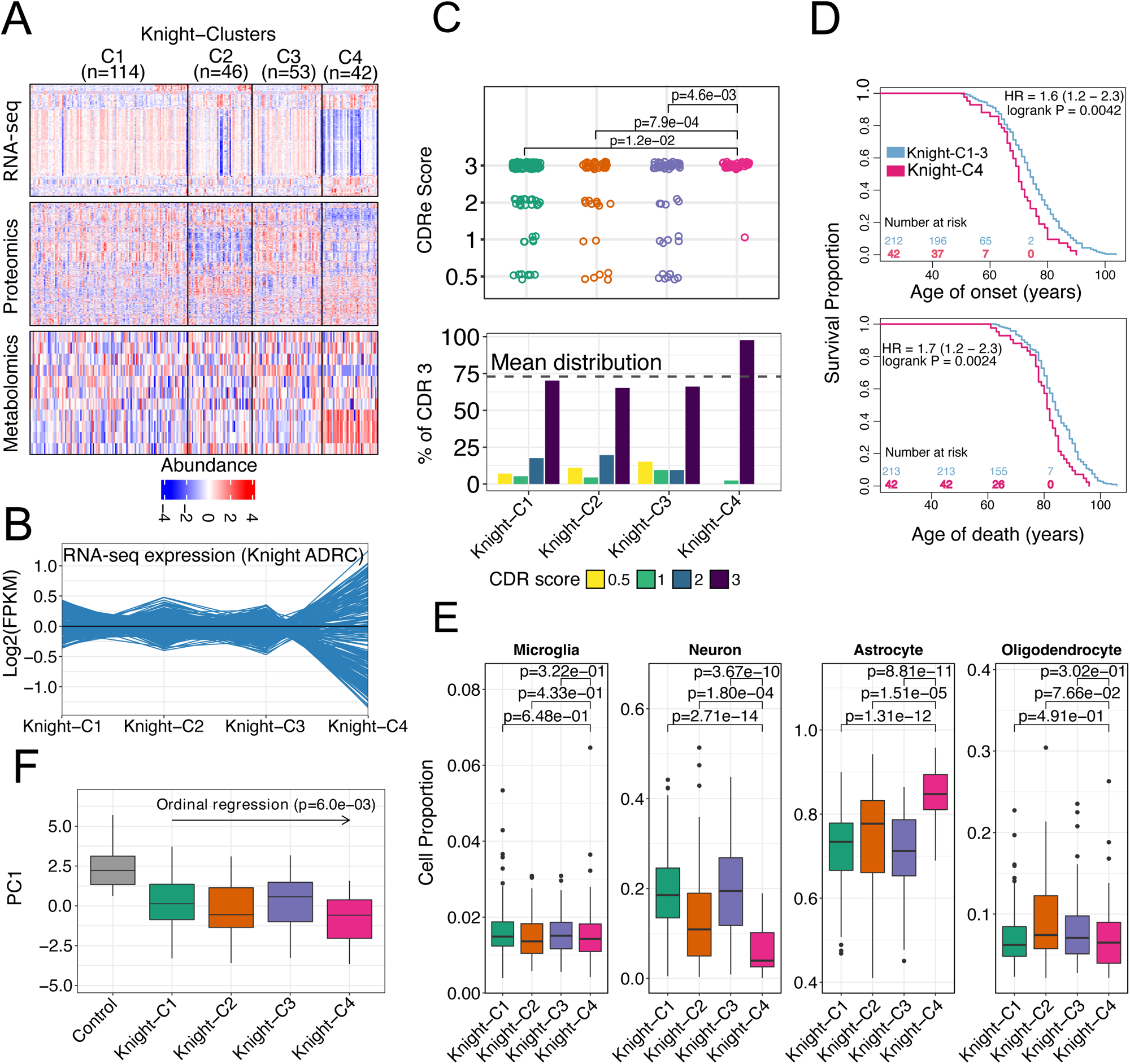
Cross-omics data integration identified four distinct molecular profiles of AD associated with worse clinical outcomes and molecular attributes. (A) Heatmaps of multi-omics profiles of the top significant features. (B) Transcriptomic profiles (scaled) of the top genes contributed significantly to the clustering, showing an overall dysregulation of brains in Knight-C4. (C) The distribution of CDR scores across four clusters with Knight-C4 associated with high CDR scores. (D) Kaplan-Meier plots showing Knight-C4 association with poor outcome for the age of onset and age of death. (E) Boxplots showing the cell proportion from deconvolution analysis across four clusters using bulk RNA-seq from four cell types. Knight-C4 is associated with a significantly higher and lower proportion of astrocytes and neurons, respectively. (F) Boxplot showing each cluster’s first principal component of metabolomics profiles, with Knight-C2 and Knight-C4 showing a lower metabolomic signature than other AD cases.

We then evaluated whether these molecular profiles were associated with genetic risk factors, neuropathological status, or clinical and demographic variables. We found that AD cases in Knight-C4 showed significantly higher Clinical Dementia Rating (CDR) scores at death (p=5.2×10^−3^; **Fig 2C**), shorter survival time following disease onset (8 months shorter, **Fig 2D**, p=4.2×10^−3^, HR=1.6) and younger age at death (**Fig 2D**, p=4.2 ×10^−3^, HR=1.7) compared to AD cases in Knight-C1-3. The Knight-C4 cluster was not associated with either AD polygenic risk score (**S2 Fig**), *APOE* ℇ4 allele carrier status (**S3A Fig**), or Braak stages (neurofibrillary tangle or amyloid) (**S3B and S3C Fig**). However, Knight-C4 included more Braak tau stage VI cases than the other clusters (**S3B Fig**). The parietal cortex is affected in later stages of AD, which could explain the lack of correlation of Knight-C4 cluster with Braak staging.

Next, we inferred the cellular population structure from bulk RNA-seq [12] leveraging digital deconvolution methods (see Methods). Knight-C4 showed significantly higher astrocyte and lower neuronal proportions (**Fig 2E** and **S2 Table**). We then evaluated whether a metabolomic profile that we previously reported to be associated with AD duration [14] would reveal additional molecular differences among these clusters. Lower values of this metabolomic profile (or *eigenmetabolite*) were associated with a longer disease duration driven by earlier onset [14]. Knight-C2 and Knight-C4 exhibited significantly lower eigenmetabolite scores (**Fig 2F**) using ordinal logistic regression (p=6.0×10^−03^). Our approach identified new molecular profiles from AD subtypes linked to cognitive test scores, the pace of disease progression, survival with the disease, the proportion of neuronal loss, astrocytosis, and metabolic changes.

### Knight-C4 profile is replicated consistently across multiple independent cohorts and brain regions

Our analyses of the transcriptomics and proteomics data from AD parahippocampal gyrus (BM36) from the MSBB study [34] (**S1 Table**) identified two distinct molecular profiles (**Fig 3A**). Cluster1 (MSBB-C1) recapitulated the transcriptomic signatures identified in Knight-C4. We also ascertained the extent of overlap between the molecules dysregulated in Knight-C4 (**S3 Table**) and MSBB-C1 (**S4 Table**) and determined a coincident profile of dysregulated genes (**Fig 3G-I**, **S4A Fig**, p < 2.2 ×10^−16^ hypergeometric test and **S5 Table**). Furthermore, MSBB-C1 also exhibited more severe dementia (p=1.5×10^−3^, **Fig 3B**) and estimated higher and lower proportions of astrocytes (p=5.5×10^−07^) and neurons (3.2×10^−09^), respectively (**Fig 3C** and **S2 Table**) compared to other AD cases in MSBB. MSBB-C1 was also associated with higher Braak tau scores (p=3.9×10^−03^, **S5A Fig**). Although not significant (p > 0.05), these donors show a trend suggesting earlier age of death (**S5B Fig**). We did not identify a significant overlap of dysregulated proteins between regions (p > 0.05, **S4B Fig**). We also observed that the molecular profile of MSBB-C1, first detected in the parahippocampal gyrus (BM36), is consistently present in multiple cortical regions, including the frontal cortex (BM10), superior temporal gyrus (BM22), and inferior frontal gyrus (BM44) (**S5C Fig**).

**Fig. 3.**
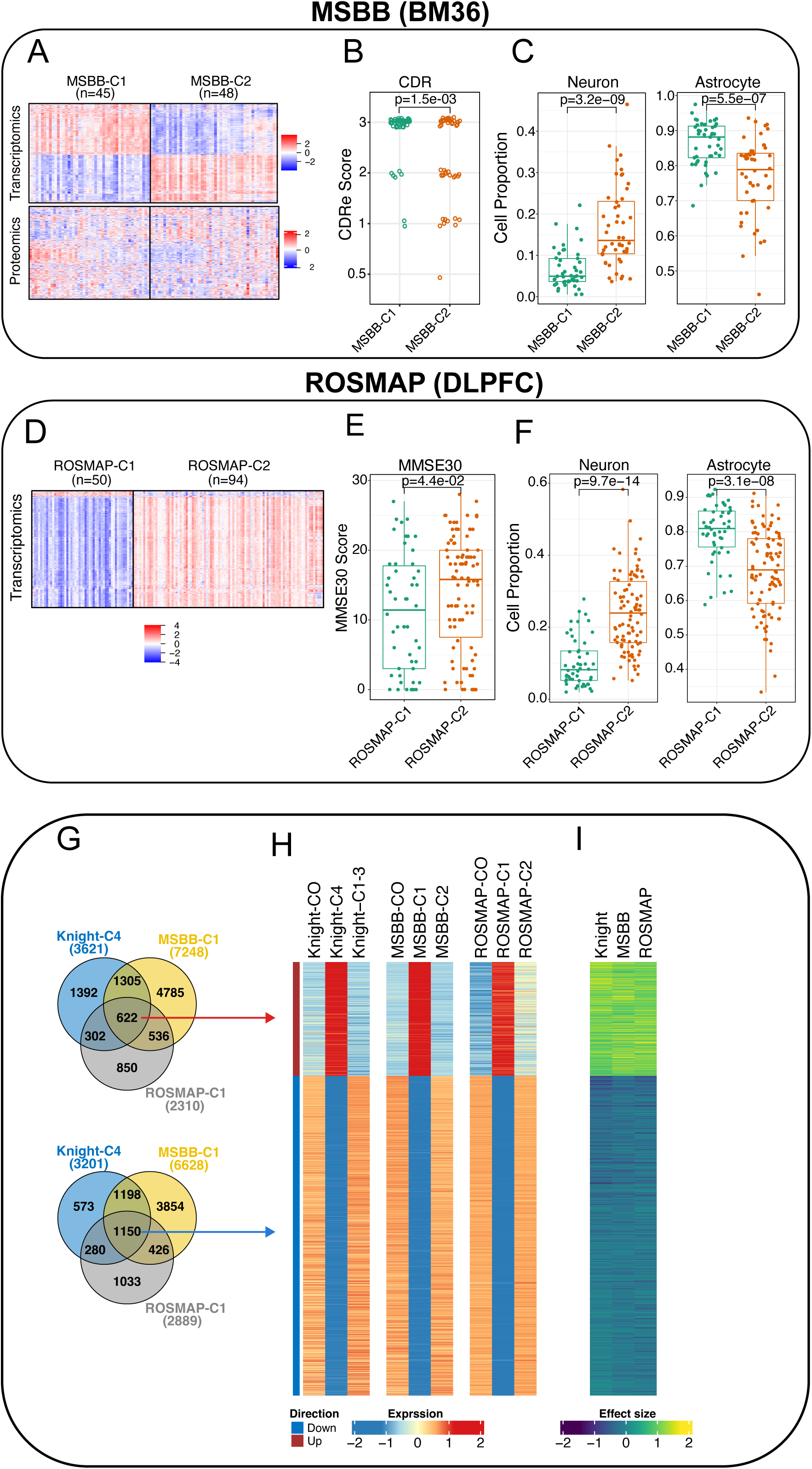
Molecular profiles of Knight-C4 are replicated in two independent datasets. (A) Heatmaps of the transcriptomic and proteomic profiles of the top features from MSBB (BM36) show two distinct clusters. (B) Boxplots showing MSBB-C1 associated with higher CDR scores, replicating Knight-C4 in the knight ADRC. (C) Boxplots showing cell proportions inferred from bulk RNA-seq from MSBB (BM36) using digital deconvolution. MSBB-C1 replicates Knight-C4 by showing an association with significantly higher and lower proportions of astrocytes and neurons, respectively. (D) Heatmap of the transcriptomic profiles of the top features from ROSMAP (DLPFC) showing two distinct clusters. (E) Boxplots showing ROSMAP-C1 associated with lower MMSE30 scores. (F) Boxplots showing cell proportion estimated from bulk RNA-seq from ROSMAP (DLPFC) using digital deconvolution. Like MSBB, ROSMAP-C1 replicates Knight-C4 by showing an association with significantly higher and lower proportions of astrocytes and neurons, respectively. (G) Venn diagram showing the common dysregulated genes in the discovery and replicated cohorts. (H) Heatmaps of the mean expression of the common genes across the discovery and replicated cohorts showing a clear, consistent expression pattern. (I) Heatmap of the effect size detected in the discovery and replicated cohorts showing a high effect size similarity across the three cohorts.

Likewise, our analyses of the RNA-seq data from the dorsolateral prefrontal cortex (DLPFC) from the ROSMAP cohort [35] identified a similar subset of AD brain donors (**Fig 3D**). Relative to those in Cluster2 (ROSMAP-C2), Cluster1 (ROSMAP-C1) recapitulated the transcriptomic profiles of those in Knight-C4 (**Fig 3G-I**, **S4C Fig,** p < 2.2 ×10^−16^ hypergeometric test and **S5 and S6 Tables**). They also showed higher tau tangle densities (p=1.3×10^−02^) and neurofibrillary tangle burdens (PHF tau tangles, p=2.1×10^−02^; **S6A and S6B Fig**), significantly higher astrocyte (p=3.1×10^−08^) and lower neuron (p=9.7×10^−14^) proportions (**Fig 3F** and **S2 Table**), and a non-significant (p > 0.05), trend to higher plaque densities—Global burden of AD pathology (**S6C Fig**, see Methods and Bennett *et al.*, 2012 [35] for details on how these neuropathological variables are identified). ROSMAP-C1 also showed evidence of greater cognitive impairment and increased neuropathological AD scores (MMSE; p=4.4×10^−02^; **Fig 3E**). Advanced AD patients with poor cognitive function show similar cellular and molecular profiles in multiple affected cortical regions. The molecular profiles associated with the severity of AD pathology can still vary even in the advanced stages of the disease.

### The Knight-C4 subjects exhibited a remarkable molecular disarray

Differential expression (DE) analyses (see Methods, **S3, S7 and S8 Tables**) showed that Knight-C4 has a ∼10-fold increase in the number of significantly differentially expressed genes (DEGs) relative to controls, in comparison to Knight-C1-C3 (**S9 Tables** and **S7A Fig**); furthermore, 90% of the 4,661 DEGs were unique to participants in the Knight-C4 (**S9 Table** and **Fig 4A-green bars**). Similarly, we observed many proteins and metabolites with significantly different abundance levels in Knight-C4 relative to controls. However, Knight-C2 has the largest dysregulated proteins (**S9 Table**, **S7B** and **S7C Fig**). We identified 24 (34%), 232 (51%), and 250 (52%) significant proteins unique to Knight-C1, 2, and 4, respectively (**Fig 4B-green bars** and **S9 Table**). We did not detect significant proteins dysregulated uniquely in Knight-C3 (**S7B Fig)**. Knight-C4 was associated with many cluster-specific metabolites significantly different from controls (90 hits; **S9 Table** and **S7C Fig**), whereas only one cluster-specific metabolite significantly different from controls was associated with Knight-C2. Of the 90 hits, 80 (90%) significant metabolites significantly differed between Knight-C4 and other AD cases.

**Fig. 4.**
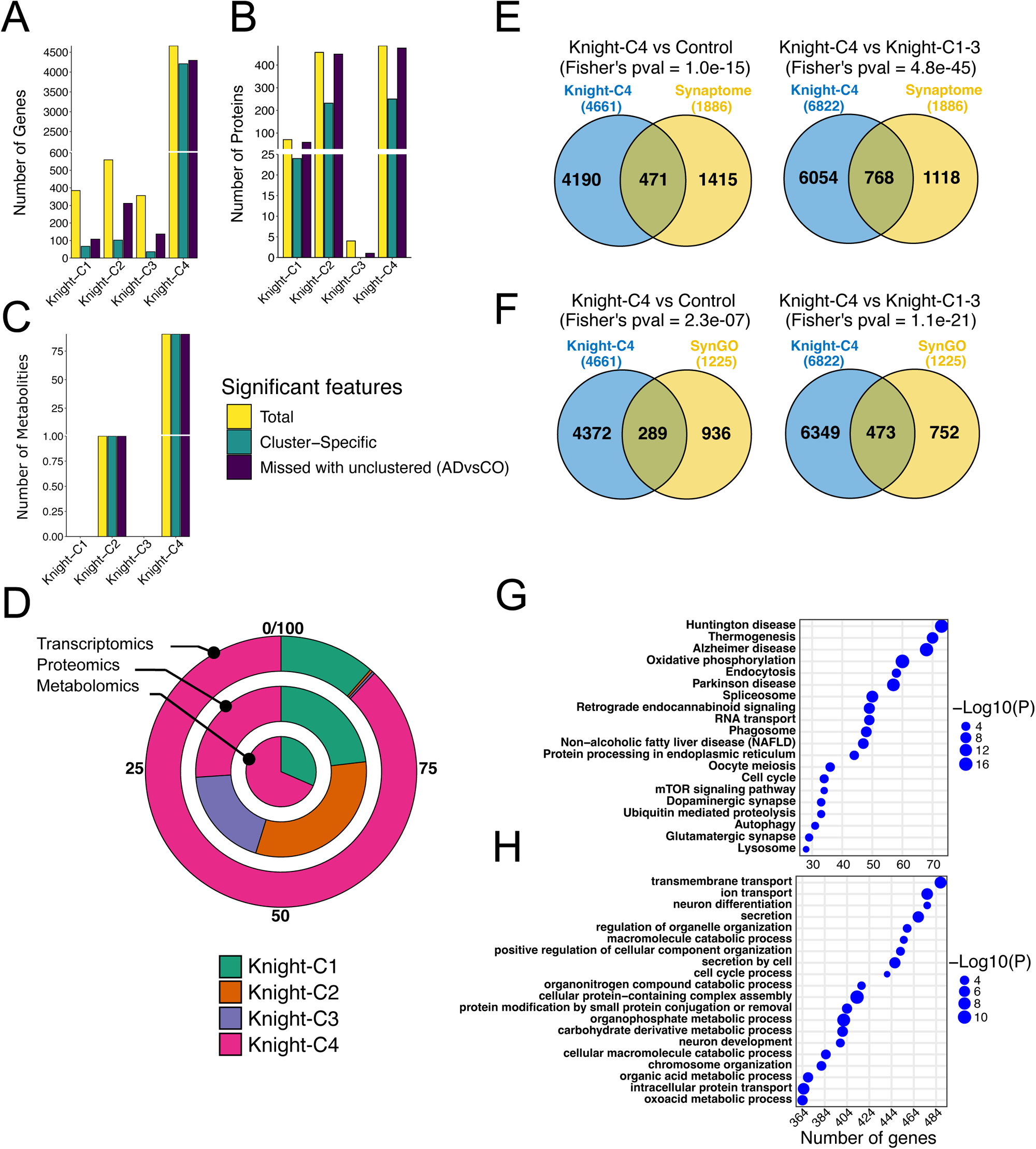
The parietal cortex of participants with worse cognitive function exhibited a remarkable molecular dysregulation. (A) Bar plots showing the significant genes detected in each cluster compared to the control. Yellow color represents the total number of genes, green represents the cluster-specific genes, and purple depicts the number of genes missed with an unclustered approach (all AD cases vs control). (B) Same as panel “a” but for proteins. (C) Same as panel “a” but for metabolites. (D) Race track plot showing the percentage of significant features for each cluster compared to other clusters for the three omics. (E) Venn diagrams showing the overlap between significant genes in Knight-C4 and synaptic genes (“synaptosome”) from Fei *et al.,* 2018^37^. (F) Same as “e” but overlapped with SynGO dataset^36^. The significance of overlap was computed using Fisher’s exact test. (G) Top 20 KEGG pathways associated with Knight-C4. (H) Top 20 GO biological process pathways associated with Knight-C4.

We also performed DE analyses comparing all AD cases combined (un-clustered) to control brains. We determined the number of significant cluster-specific features not captured by unclustered analyses (**S9 Table** and **Fig 4A-4C-purple bars**). We observed that Knight-C4 exhibited the highest number of features missed by un-clustered analyses with 4,295 genes (92%), 475 proteins (98%), and 90 metabolites (100%) (**S9 Table** and **Fig 4A-4C-purple bars**), indicating the importance of clustered analyses. Notably, a high percentage of features was also observed with Knight-C2 in proteomics (98%). In contrast, un-clustered analyses identified a small percentage of significant features not captured by clustered analyses. For instance, only 5% and 0.5% of un-clustered significant features for transcriptomics and proteomics, respectively, were not captured by Knight-C4. Knight-C4 missed no un-clustered significant metabolites.

DE analyses comparing brains in each AD cluster to brains in other AD clusters (**S3, S7 and S8 Tables**) revealed that Knight-C4 showed the highest percentage of significant hits compared to other AD brains (**Fig 4D** and **S10 Table**). This significant difference was observed in transcriptomics, with 6,822 significant hits compared to 678, 22, and 1 for Knight-C1-3 (**Fig 4D-outer track** and **S10 Table**). Similarly, this difference was observed in metabolites with 228 significant hits in Knight-C4 compared to 105, 0, and 0 for Knight-C1-3, respectively (**Fig 4D-inner track** and **S10 Table**). No significant differences in the number of hits were observed between the four clusters in proteomics, with all clusters showing relatively similar detections (**Fig 4D-middle track** and **S10 Table**). We also verified that none of the omics dominate this solution, each capturing molecular heterogeneity among AD brains that are not necessarily detected by other *omics* modalities.

Noticeably, these DE analyses also showed that Knight-C4 is significantly enriched in dysregulated synaptic genes (**Fig 4E** and **4F** and **S11 Table**) using the annotation from The SynGO consortium [36] (p=1.0×10^−15^ and p=4.8×10^−45^ for Knight-C4 vs. Control and Knight-C4 vs. Knight-C1-3 sets respectively) and curated list from Fei *et al.,* 2018 [37] (p=2.3×10^−07^ and p=1.1×10^−21^ for Knight-C4 vs. Control and Knight-C4 vs. Knight-C1-3 sets respectively). MSBB-C1 and ROSMAP-C1 profiles were also significantly enriched in synaptic genes (**S8A** and **S8B Fig** and **S11 Table**).

In summary, these results show that molecular profiles of AD are heterogeneous, and the cross-omics integration approach can better identify the substantial differences between clinically relevant sub-categories of AD brains.

### The Knight-C4 cluster is significantly enriched in AD-related dysregulated pathways

Knight-C4 contains multiple pathways related to different aspects of neurodegeneration (**Fig 4G** and **4H**). For instance, KEGG pathways identified multiple intracellular trafficking and signaling pathways (**Fig 4G**), including (e.g., *VPS29* and *VPS35*) endocytosis (p=6.5×10^−04^), (e.g., *ATP6V1A* and *RAC1*) phagosome (p=1.2×10^−06^), and (e.g., *GSK3B*, and *IGF1)* mTOR signaling pathway (p=3.3×10^−02^), all of which have been shown to play major roles in the pathological processes of AD [38–40]. Genes (*PPT1*, *GBA*, *HEXB*, and *LGMN)* from lysosomal pathway, which is critical for microglia, and neurons, were also identified (p=4.7×10^−02^, **Fig 4G**). Growing evidence has shown that the dysfunction of lysosomes plays a major role in the pathogenesis of multiple neurodegenerative diseases (ND), including the progression of AD [41–45]. Knight-C4 was also associated with genes (*BECN1* and *MAPK1)* from the autophagy pathway (p=1.6×10^−02^, **Fig 4G**), which is associated with several neurodegenerative disorders, including AD [46–49]. For instance, autophagy dysfunction was shown to be linked to the accumulation of misfolded protein aggregates [50]. The downregulation of Knight-C4 compared to other AD cases identified multiple ND-related pathways, including AD (p=2.5×10^−14^), Huntington disease (p=3.3×10^−14^), and PD (p=2.1×10^−12^) (**Fig 4G**), Knight-C4 also exhibited downregulation of synaptic pathways including dopaminergic synapse (p=7.0×10^−03^) and glutamatergic synapse (p=1.0×10^−02^) (**Fig 4G**), including *GRIA1*, *GRIA2, PPP3CA*, *GRIN2A*, *PLCB1*, *CALY*, and *GLS* genes. Furthermore, GO biological process analyses identified significant neuronal differentiation (p=1.8×10^−02^) and development (p=5.7×10^−03^) associated with Knight-C4 (**Fig 4H**). Most of these pathways were also identified in MSBB-C1 and ROSMAP-C1 (**S9A-S9C Fig**).

Compared to the control group, Knight-C4 showed additional pathways, including herpes simplex virus 1 infection (p=4.4×10^−03^), a risk factor for AD [51–54] that lead to neuronal damage and induced gliosis [55,56], and synaptic vesicle cycle (p=4.6×10^−05^) shown to play a key role in the pathobiology of AD [57] (**S9D Fig**). Noticeably, herpes simplex virus 1 infection was significantly enriched in Zinc Finger genes, a class of protein-coding genes known to regulate gene expression at the transcriptional and translational levels by binding to bind to certain DNA or RNA molecules. GO biological process analyses identified several significant synaptic signaling and transmission pathways including two well-known AD biomarkers, *SNAP25* and *SYT1*. (**S9E Fig**). Many of these pathways were also present in MSBB-C1 and ROSMAP-C1 (**S10A-S10D Fig**). Pathway analyses using significant proteins identified AD-related pathways including PI3K−Akt signaling (p=6.5×10^−19^), rap1 signaling (p=5.8×10^−14^), axon guidance (p=2.1×10^−12^), MAPK signaling (p=1.4×10^−20^), and ras signaling (p=1.8×10^−15^) pathways associated with the downregulation of Knight-C4 (**S10E Fig**). Noticeably, *MAPT*, the gene that encodes the microtubule-associated protein tau (MAPT), was among the genes in the MAPK signaling pathway. Top GO pathways, enriched in Knight-C4 downregulated proteins (and replicated in MSBB-C1), were related to cell/gene regulatory pathways (**S10F Fig**). Through cross-omics analyses, we discovered numerous known and novel pathways contributing to the cognitive decline in AD patients in multimodal profiles.

### The multimodal clusters identify potential AD biomarkers

Among the top genes/proteins dysregulated in Knight-C4, several AD biomarkers including *APP, APOE, CLU, SNAP25, GFAP, SNCA, NOTCH3, TARDBP, GRN, MMP9,* and *C9ORF72* were identified (**S3, S7**, **S8 Tables** and **Fig 5A** and **5D**). Of these, transcript levels of Synaptosome Associated Protein 25 (*SNAP25*), a protein implicated in AD and Down syndrome (DS) [58], were significantly downregulated in Knight-C4 (**Fig 5B**), as it has been previously reported in AD [58]. *SNAP25* protein levels were also shown to be reduced in the CSF in AD patients [59]. *SNAP25* was significantly DE between Knight-C4 and control (p=5.9×10^−3^) and between Knight-C4 and other AD cases (p=4.2×10^−08^, **Fig 5B**). However, *SNAP25* was not significantly DE (p > 0.05, **Fig 5B**) in non-clustered AD cases compared to the control. No significant differences were observed in *SNAP25* protein levels in clustered or un-clustered analyses. The decreased expression of *SNAP25* in Knight-C4 was replicated in MSBB-C1 (transcript and protein levels) and ROSMAP-C1 (transcript levels) cohorts (**S11A-S11C Fig**).

**Fig. 5.**
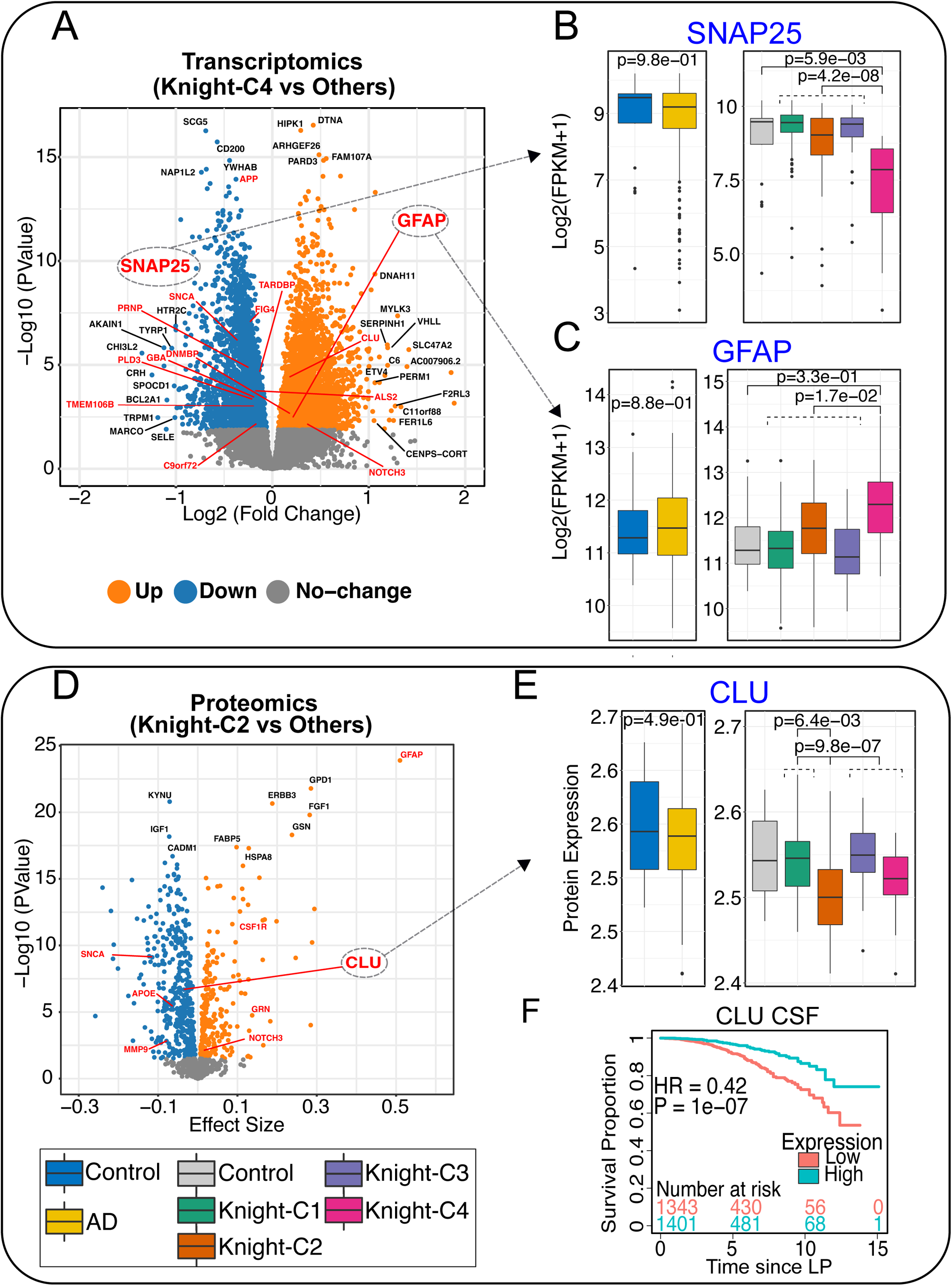
Differential expression analyses identified neurodegenerative disease genes associated with distinct clusters. (A) Volcano plot showing the up-and downregulated genes identified between Knight-C4 vs other clusters. Genes in red are examples of known neurodegenerative disease genes. Genes in black are the top 10 significant genes based on the adjusted p-value and genes with log2(fold change) > 1. (B) Boxplots showing the transcriptomic profiles (in FPKM) of *SNAP25* across four clusters (right) and for AD cases combined (left) in the Knight ADRC cohort. (C) Same as “b” but for *GFAP*. (D) Volcano plot showing the up-and downregulated proteins identified between Knight-C2 vs other clusters. (E) Boxplots showing the proteomic profiles of *CLU* across four clusters (right) and for all AD cases (left) in the Knight ADRC cohort. (F) Kaplan-Meier plot showing the association of Clusterin (ApoJ) protein concentrations in CSF with an increased risk of dementia progression using CDR change from 0 to CDR > 0 using the CSF dataset from the Knight ADRC participants.

Glial Fibrillary Acidic Protein (*GFAP*), an emerging fluid biomarker in brain disorders [60] usually employed to identify astrocytic reactivity state, a well-documented process in AD and neurodegeneration, was identified. The increased astrocyte reactivity and overexpression of *GFAP* have been reported in multiple studies [58,60–68]. *GFAP* was overexpressed uniquely in Knight-C4 compared to Knight-C1-3 (**Fig 5C**, p=1.7×10^−2^). However, *GFAP* was not significantly DE in Knight-C4 compared to controls, nor was it DE in all AD cases compared to the control (p > 0.05, **Fig 5C**). We repeated these analyses using a simplified model that did not correct for cellular population structure to examine whether collinearity between case-control status and cellular population structure may introduce spurious results, but we could not obtain a significant association. The same pattern was observed with *AQP4*, another marker gene of astrocyte reactivity state; it was only significantly different between Knight-C4 and Knight-C1-3 (p=1.0×10^−02^). We did not identify significant differences in *GFAP* protein levels (p > 0.05, **S12 Fig**). However, protein levels of *GFAP* were significantly upregulated in Knight-C2 (**S12 Fig**) compared to the control and AD brains in Knight-C1,3,4 (p=1.0×10^−4^ and 1.4×10^−21,^ respectively). The increased transcriptomic *GFAP* levels were also identified in MSBB-C1 and ROSMAP-C1 (**S13A-S13C Fig**) but with less significance level in ROSMAP-C1. Altogether these results support heterogeneity in the presence of astrocytic reactivity state and its association with different AD subtypes.

Our cross-omics integration analyses also identified cluster-specific dysregulation of Clusterin (*CLU*), a gene associated with AD [69,70]. Genome-wide association studies have shown that *CLU* (*APOJ*) is the third most associated late-onset AD (LOAD) risk gene after *APOE* and *BIN1* [70–73]. It was also shown to play a neuroprotective role in AD by altering Aβ aggregations and promoting Aβ clearance [73–77]. *CLU* protein levels were significantly downregulated in Knight-C2 compared to the control (p=6.4×10^−3^) and AD cases in Knight-C1,3,4 (p= 9.8×10^−7^; **Fig 5E**). *CLU* was also significantly downregulated in Knight-C4 compared to the control (p=3.4×10^−02^; **Fig 5E**). We did not observe significant differences in *CLU* transcriptomic and proteomic levels in the MSBB and ROSMAP cohorts. To assess the clinical implications of *CLU* dysregulation, we evaluated its levels in the CSF from 553 participants of the Knight ADRC by performing survival analyses for progression using CDR change from 0 to 0.5. These analyses indicate that lower CSF *CLU* is associated with an increased risk of dementia progression (HR=0.42; p=1.0×10^−07^, **Fig 5F**). Together, results show that cross-omics approach identified potential biomarkers for AD that traditional approaches may miss.

### Metabolic differential abundance and pathway analyses in Knight-C4

We next looked at the significant metabolites that uniquely characterize brain samples in Knight-C4. To determine whether significant metabolites are replicated in additional cohorts, we accessed metabolomics from the same ROSMAP cohort [35] (**S1 Table**, Methods). We performed differential abundance analyses comparing the levels from donors in the previously identified ROSMAP-C1 cluster to controls and other AD cases (**S6 Table**). We identified 57 significant metabolites (43 increased and 14 decreased) in Knight-C4 compared to controls replicated in the ROSMAP-C1 (**Fig 6A** and **S12 Table**). Similarly, we identified 74 metabolites (70 increased and 4 decreased) in the Knight-C4 compared to other AD cases replicated in ROSMAP-C1 (**Fig 6B** and **S12 Table**).

**Fig. 6.**
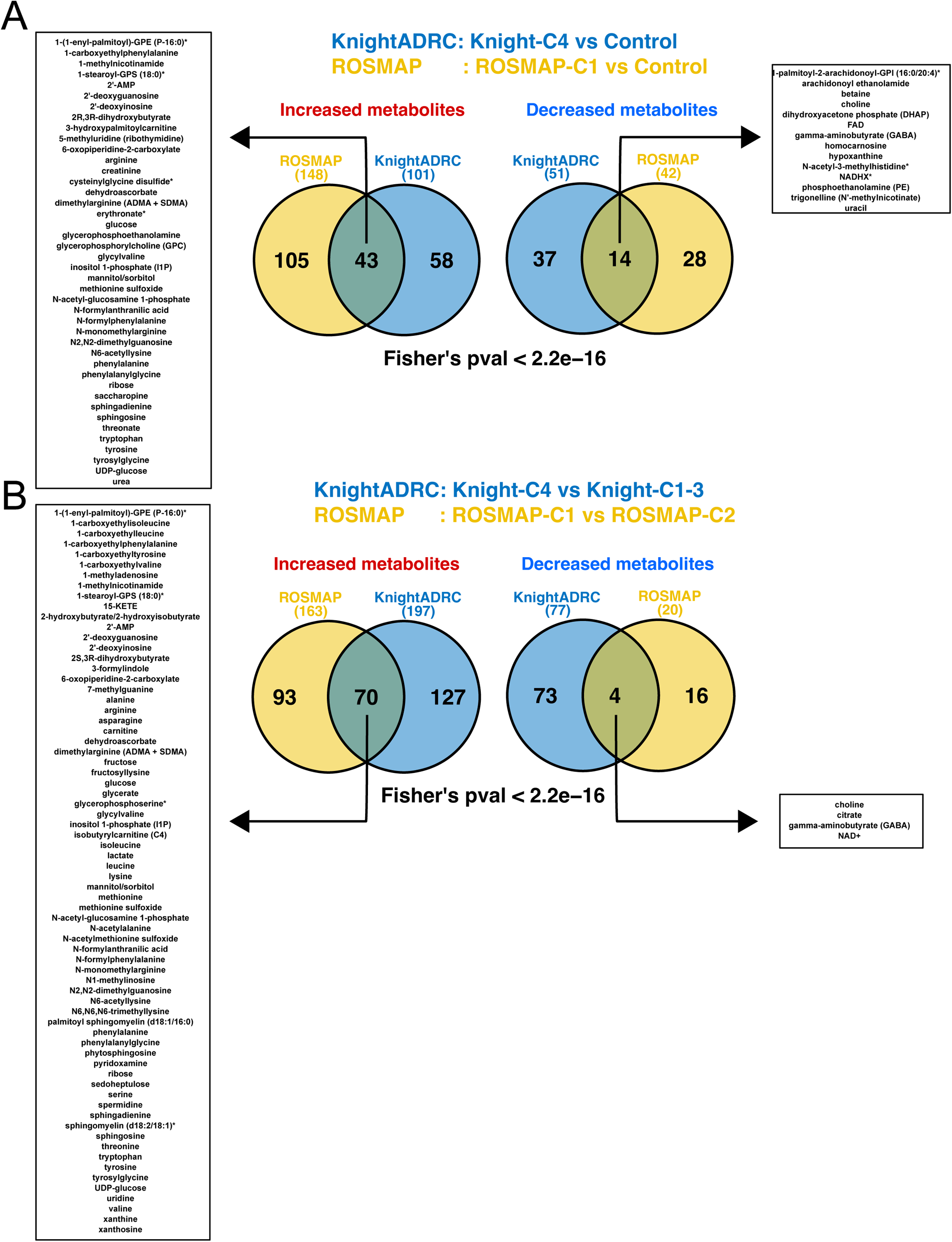
Differential abundance analyses identified significant metabolites and pathways associated with Knight-C4. (A) Venn diagrams show the significantly increased and decreased metabolites shared between the Knight-C4 and ROSMAP-C1 compared to the control. The left and right boxes show the names of shared metabolites. (B) Similar to panel “a” but for metabolites shared between Knight-C4 and ROSMAP-C1 compared to the other cases. The significance of overlap was computed using Fisher’s exact test.

Pathway analyses using these common metabolites identified several pathways significantly related to AD. Among the metabolites decreased in AD (**Fig 6A** and **6B**), gamma-aminobutyrate (GABA) and choline were identified, which involved major neurotransmitter pathways. GABA, synthesized in neurons, was significantly decreased in Knight-C4 (p= 5.1×10^−04^) and ROSMAP-C1 (p=1.3×10^−02^) compared to the control (**S14A Fig**). The same observation was found when these profiles were compared to other AD cases (p=1.2×10^−05^ and p= 1.6×10^−02^ for the Knight-C4 and ROSMAP-C1, respectively, **S14A Fig**). As an inhibitory neurotransmitter, GABA has been shown to play a key role in synchronizing the activity of the human cerebral cortex [78,79]. Similarly, the metabolomics profiles of choline were decreased in these profiles in both Knight-C4 (p= 4.1×10^−06^) and ROSMAP-C1 (p=1.2×10^−04^) compared to controls (**S14B Fig**). These reading levels of choline also show significantly different abundance in these clusters compared to other AD cases (p= 2.9×10^−07^ and p=4.7×10^−3^ for Knight-C4 and ROSMAP-C1, respectively, **S14B Fig**). Choline was also shown to have a critical role in neurotransmitter function because of its effect on the function of acetylcholine and dopamine [80]. However, we did not find significant differences in the reading levels of these active neurotransmitters. Nor did we observe significant differences for additional neurotransmitters such as serotonin and glutamate. We then studied tryptophan, an amino acid essential for metabolites that serve as neurotransmitters and signaling molecules [81]. We found that its levels increased in the Knight-C4 compared to the control and other AD cases (p=8.3×10^−3^ and 3.0×10^−06,^ respectively) (**S15A Fig**). Similarly, we found that brains in ROSMAP-C1 showed increased tryptophan levels compared to the control and other AD brains (p=1.4×10^−4^ and 2.8×10^−07,^ respectively) (**S15A Fig**).

Several increased carbohydrate/sugar metabolites were identified in Knight-C4 (**Fig 6A** and **6B**), including glucose, mannitol, sorbitol, ribose, and UDP-glucose. For example, glucose, a metabolite implicated in the pathogenesis of AD [82,83], was significantly increased in Knight-C4 and ROSMAP-C1 (**S15B Fig**). Components of glycolysis and the tricarboxylic acid (TCA) cycle were also dysregulated. Specifically, levels of citrate were decreased in both Knight-C4 (p= 9.7×10^−05^) and ROSMAP-C1 (p= 1.4×10^−03^) compared to other AD cases (**S16A Fig**). NAD+, an important oxidizing cofactor in glucose metabolism and cellular respiration, was also decreased in both Knight-C4 (p=2.2×10^−02^) and ROSMAP-C1 (p=2.2×10^−02^) compared to other AD cases (**S16B Fig**). NAD+ is synthesized from tryptophan, and levels of NAD+ have been shown to decrease with age, potentially contributing to the pathogenesis of AD [84]. These observations align with previous findings that associate increased brain glucose and slowed glucose metabolism with more severe AD [83,85]. In addition, several increased sphingolipid metabolites were identified, including sphingomyelin, sphingosine, and sphingadienine (**Fig 6A** and **6B**). Previous studies showed the implication of sphingolipid metabolism deregulation in AD [86]. For instance, sphingosine, which had higher levels in AD brains [86], showed significantly increased metabolism levels in Knight-C4 and ROSMAP-C1 (**S16C Fig**). Altogether, these results show that cross-omics methods can uncover dysregulated metabolites in AD brains, indicating a link to other conditions like diabetes [85].

### Integration with single-cell transcriptomic data reveals cell type-specific changes associated with distinct molecular profiles of AD

We studied the cell type-specific pathways altered in the AD profiles by leveraging single-nuclei RNA-seq from a subset of the Knight ADRC participants. We used the top 5% of genes that showed the most specific cell-type expression profile in neurons, astrocytes, oligodendrocytes, oligodendrocyte precursor cells (OPC), and microglia. Then, we used this information to determine if any AD molecular profiles showed a stronger signature of cell-type specific dysregulation. These analyses showed that Knight-C4 was enriched in genes overexpressed in astrocytes and endothelial cells (38% and 23% for all hits, respectively; **Fig 7A-left panel** and **S13 Table**). Whereas the first may indicate reactive astrogliosis, the latter may suggest endothelial dysfunction in AD development, providing more evidence of vascular involvement in this disease. Pathways analyses of astrocytic genes identified astrocyte-related pathways (**Fig 7B**), including axon guidance (p=2.8×10^−05^) and hedgehog signaling pathway (p=1.6×10^−06^). Previous work has shown the role of astrocytes in axon guidance during development and repair [87] and hedgehog signaling pathways involved in neuroinflammation, neuronal cell differentiation, and neuronal death [88]. Examples of genes involved in these pathways are *FYN*, *SMO*, *KIF7*, and *GLI2*. Many of these pathways were also enriched in overexpressed genes in endothelial cells (**S17A Fig**).

**Fig. 7.**
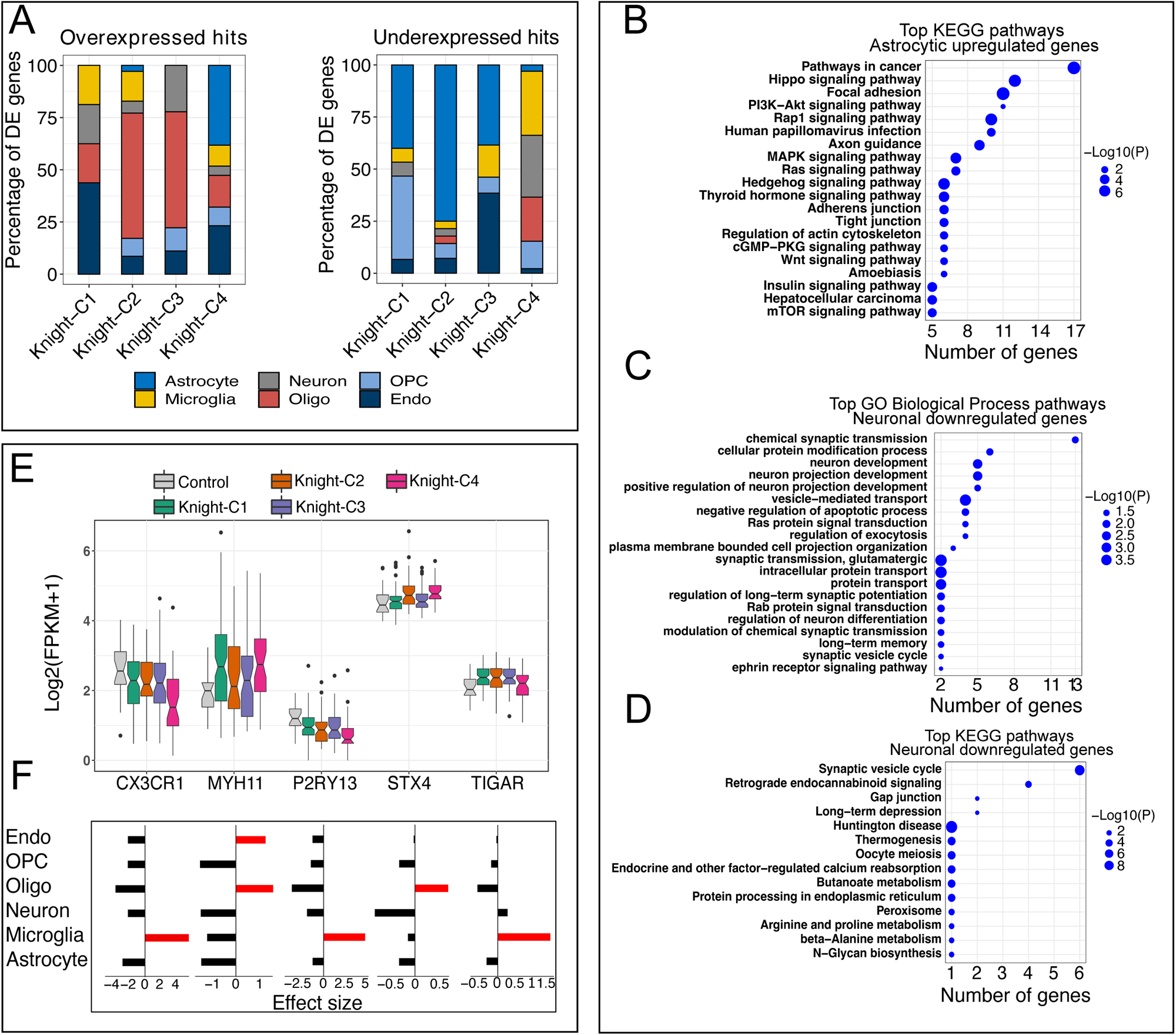
Integrating single-nuclei data with cross-omics profiles identified cell-type-specific genes and pathways. (A) Bar plots showing the percentage of up-and downregulated genes (clusters vs. control) identified in each cell type. (B) Top 20 KEGG pathways enriched in astrocytic upregulated genes. (C) Top 20 GO biological process pathways enriched in neuronal downregulated genes. (D) Top KEGG pathways enriched in neuronal downregulated genes. (E) Boxplots show the transcriptomic profiles (in FPKM) of examples of cell-type specific genes identified in the Knight ADRC cohort. (F) The effect size of these examples was generated from single-nuclei data from the Knight ADRC participants. Red bars represent the cell type in which the gene is overexpressed.

Knight-C4 has also enriched in genes downregulated in microglial and neuronal genes (**Fig 7A-right panel** and **S13 Table**). Neuronal genes showed an association with synaptic vesicle cycle (p=1.0×10^−06^) and retrograde endocannabinoid signaling (p=3.6×10^−03^) [89–91], and gap-junction (p=2.2×10^−02^) formation known to play an essential role in intercellular metabolic communication and transmission across electrical synapses [92] (**Fig 7C** and **7D**). Two AD biomarker genes, *SNAP25* and *SYT1,* were included in these pathways. Microglial downregulated genes implicate AD-related pathways, including phagosome (p=1.0×10^−06^), herpes simplex virus 1 infection (p=2.0×10^−04^)—previously associated with AD risk [51–54], gap junction (p=2.2×10^−02^), and AD (p=5.4×10^−08^) (**S17B Fig**). Two interesting examples of genes represented in these pathways are *C3* and *CD74*, which are implicated in AD pathology. Further explorations showed that Knight-C4 is significantly enriched in downregulated genes related to homeostatic microglia, including C-X3-C Motif Chemokine Receptor 1 (*CX3CR1;* p= 5.10x^−03^, **Fig 7E** and **7F**) and purinergic receptor P2Y13 (*P2RY13;* p=1.2×10^−2^, **Fig 7E** and **7F**).

Additionally, we identified genes dysregulated in specific clusters and cell types, including the TP53-induced glycolysis regulatory phosphatase (*TIGAR*) overexpressed in microglia and upregulated in Knight-C1 (**Fig 7E** and **7F**). *TIGAR* is a p53-inducible protein that regulates energy metabolism and oxidative stress. Among the upregulated endothelial-specific genes in Knight-C1, we identified *MYOCD*, *CNN1*, *PLN*, *MYH11*, S*100A4*, *SLC13A4*, *DES*, implicating pathways associated with cell cycle, junction, anion transport, Aβ clearance and actin. In particular, *MYH11* overexpressed in Knight-C4 (**Fig 7E** and **7F**) has been reported to be associated with the risk of dementia [93]. Moreover, oligodendrocyte-specific genes dysregulated in Knight-C2 include *STX4* (Syntaxin-4), *ACTN4*, and *FBXO32* relating to pathways associated with vesicle docking, neural tube, junction, apoptosis, and actin. Of particular interest, *STX4* upregulated in Knight-C2,4 (**Fig 7E** and **7F**) is critical for vesicle docking and the Wnt frizzled receptor (*FZD9*) and coreceptor (*LRP5*), regulating the synaptic vesicle cycle [30]. *STX4* has also been shown as one of the disease risk genes in AD [94].

These results suggest that combining cross-omics data with single-cell resolution allows uncovering of molecular profiles associated with AD features to unprecedented granularity and complexity.

### *SNCA* is a top hit in the multimodal cluster Knight-C4

Transcriptomic and proteomic assays showed *SNCA* among the top gene/proteins that best discriminate Knight-C4 versus Knight-C1-3 molecular profiles. *SNCA* transcriptomic levels were significantly downregulated in Knight-C4 compared to Knight-C1-3 (p= 2.0×10^−05^) and control brains (p=2.2×10^−02^; **Fig 8A**). Interestingly, we did not observe a significant association (p > 0.05) in non-clustered AD compared to the control (**Fig 8A-left panel**). *SNCA* encodes alpha-synuclein (αSyn) protein, whose protein levels were marginally downregulated in brains from Knight-C4 (**Fig 8B**) compared to controls (p=5.16×10^−02^) and significantly lower in Knight-C2 compared to controls (p=7.8×10^−5^) and Knight-C1,3,4 (p=6.2×10^−09^).

**Fig. 8.**
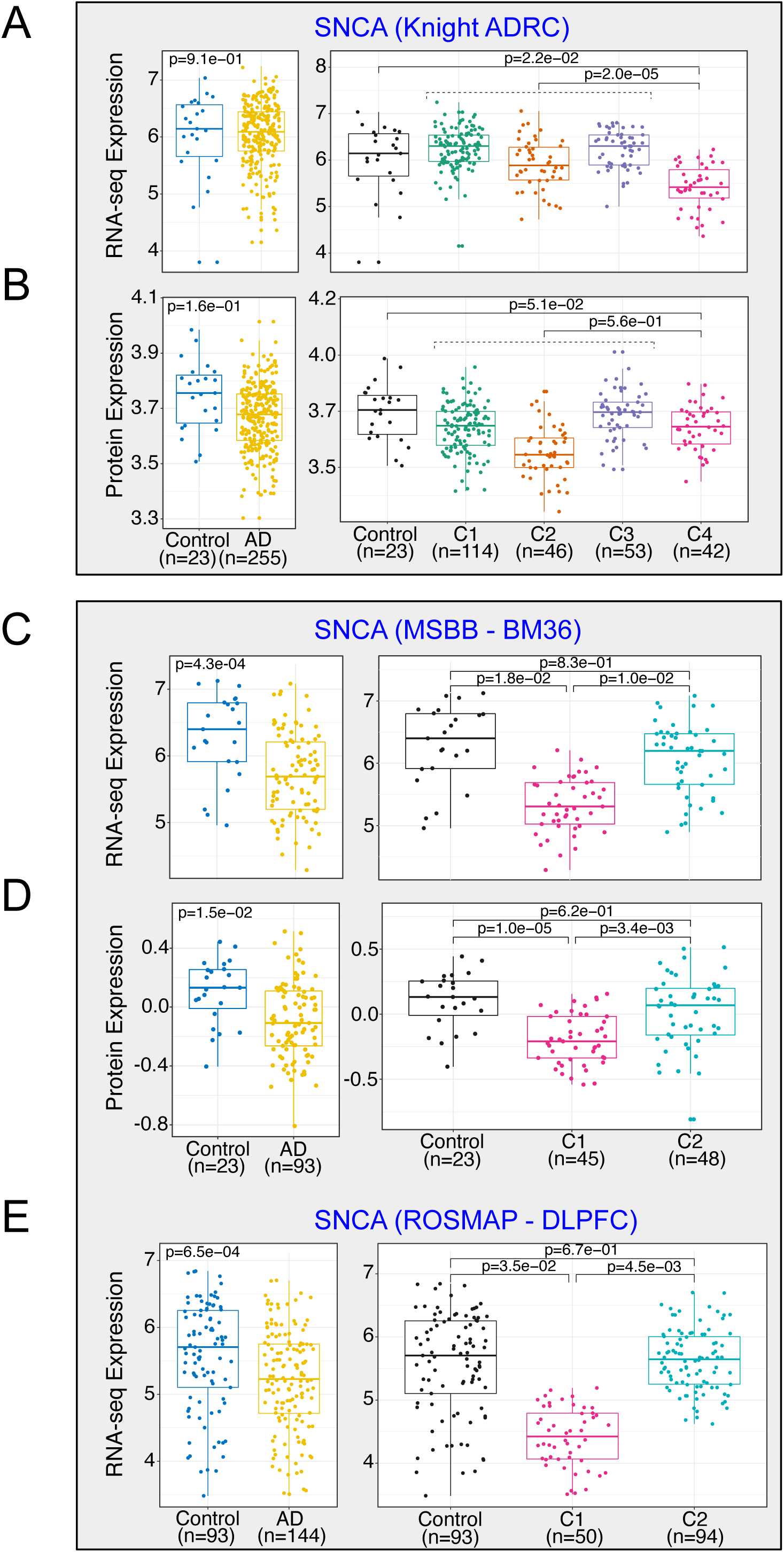
Cross-omics integration identified alpha-synuclein levels downregulated in AD participants with worse cognitive function common across multiple brain regions. (A) Transcriptomic profiles (in FPKM) of *SNCA* across four clusters (right) and for all AD cases (left) in the Knight ADRC cohort. (B) Proteomic profiles of *SNCA* across four clusters (right) and all AD cases (left) in the Knight ADRC cohort. (C) Transcriptomic profiles of *SNCA* across four clusters (right) and all AD cases (left) in the MSBB BM36 cohort. (D) Proteomic (TMT) profiles of *SNCA* across four clusters (right) and all AD cases (left) in the MSBB BM36 cohort. (E) Transcriptomic profiles of *SNCA* across four clusters (right) and all AD cases (left) in ROSMAP DLPFC cohort.

*SNCA*/*αSyn* is also downregulated in MSBB-C1 (**Fig 8C** and **8D**) compared to the controls (p=1.8×10^−02^ and p=1.0×10^−05^ for transcriptomics and proteomics, respectively). Similarly, *SNCA* levels in MSBB-C1 were significantly lower than other AD (p=1.0×10^−02^ and p=3.4×10^−03^ for transcriptomics and proteomics, respectively, **Fig 8C** and **8D**). However, no significant differences in transcript or protein levels of *SNCA* were observed in MSBB-C2 compared to controls (p > 0.05, **Fig 8C** and **8D**). Downregulation of *SNCA* transcript in the MSBB-C1 donors was also seen in the frontal pole (BM10), superior temporal gyrus (BM22), and inferior frontal gyrus (BM44) brain regions (**S18A-S18C Fig**). Similarly, *SNCA* transcriptomic levels were significantly downregulated in ROSMAP-C1 (**Fig 8E**) compared to the control (p=3.5×10^−02^) and other AD cases (p=4.5×10^−03^).

*αSyn* is the major constituent of Lewy bodies, the histopathologic hallmark of PD and DLB. LB pathology is also present in most advanced AD cases, including autosomal dominant AD and DS [95–101]. In addition, previous work has demonstrated the relationship between *αSyn* and AD pathologies, including Aβ plaques and tau aggregations using different mouse models [102,103] and implicated *αSyn* pathology in neuronal/synaptic dysfunction and death [104,105]. Co-pathology with *αSyn* is present in 50-60% of AD brains [95,96,106]. However, it is not clear if it plays any role in AD pathophysiology. In animal models, it has been shown that adding *αSyn* to an amyloidosis model increased neuron loss that correlated with the progressive decline of cognitive and motor performance, and a “feed-forward” mechanism whereby Aβ plaques enhance endogenous α-syn seeding and spreading over time has been propose^87^. Mutations in *SNCA*, overexpression of wildtype *αSyn*, and injection of pre-formed *aSyn* fibrils (PFFs) are sufficient to cause neuropathology and parkinsonian syndromes in both humans and animal models [107–113].

We also analyzed NanoString gene expression data (see Methods) generated from a panel of 807 gene transcripts measured in brain tissue of A53T *αSyn* transgenic mice crossed onto *APOE* backgrounds [114]. The *SNCA* A53T mutation cause familial Parkinson’s disease. Of the 807 genes included in the panel, 89 (11%) were classified as synaptic genes (SynGO consortium [36]). We performed DE analysis to compare mice with high pSyn pathology (defined as > 5% coverage by IHC) to mice with low pathology (< 5% coverage by IHC, see Methods). We identified 255 DEGs, including 13 synaptic genes. We then interrogated which synaptic genes in DE in Knight-C4 were also dysregulated in the A53T αSyn-APOE transgenic mice (**S19 Fig**). We identified that all 13 synaptic genes identified as dysregulated in the mouse model were also dysregulated in Knight-C4 compared with other AD cases (Fisher p=3.6×10^−09^) (**S19A Fig**). These results suggest that the profile of Knight-C4 is reproducible in mouse models of aggregation and neurodegeneration. Compared to the control, only five synaptic genes were shared between the three datasets (**S19B Fig**). These findings were limited because the nanoString panel contains only 12% of the significant genes in Knight-C4. These results show that integrating data from multimodal clusters in humans with molecular data from an established mouse model of neurodegeneration due to *αSyn* provides new clues about the role of known proteins in AD pathophysiology.

### Cross-omics integration identified CSF synaptic biomarkers for the molecular staging of AD

Knight-C4 exhibits significant dysregulation of synaptic genes and association with AD pathology but critically, is also associated with poor cognitive performance. Thus, we hypothesized that synaptic changes associated with AD molecular multimodal profiles could reflect multiple stages of AD. To evaluate this, we selected those genes dysregulated in Knight-C4 and ROSMAP-C1, and examined the effect size differences between the early-AD vs. control compared to that of the late-AD vs. control (**Fig 9A**). Interestingly, we observed a concordant change of synaptic gene dysregulation in early- and late-AD compared to controls; however, the effect was more pronounced at later stages. (**Fig 9A and 9B**, R^2^ = 0.96, p < 2.2×10^−16^, slope =0.75). This was consistent among 170 synaptic genes, including *IGF1*, *NRXN3*, and *YWHAZ* (**Fig 9C**). Co-expression analyses (see Methods) identified a core module (**S20 Fig, turquoise color**) highly enriched in interacting proteins, including *SNAP25*, *APP*, and *SNCA* (**Fig 9D**). Subsequent pathway analyses identified several AD-related pathways, including 160 protein-protein interaction hub proteins (**Fig 9E** and **S14 Table**). Additional gene interaction analyses showed that those proteins have a strong interaction, including physical and genetic interactions (**Fig 9F-9H**).

**Fig. 9.**
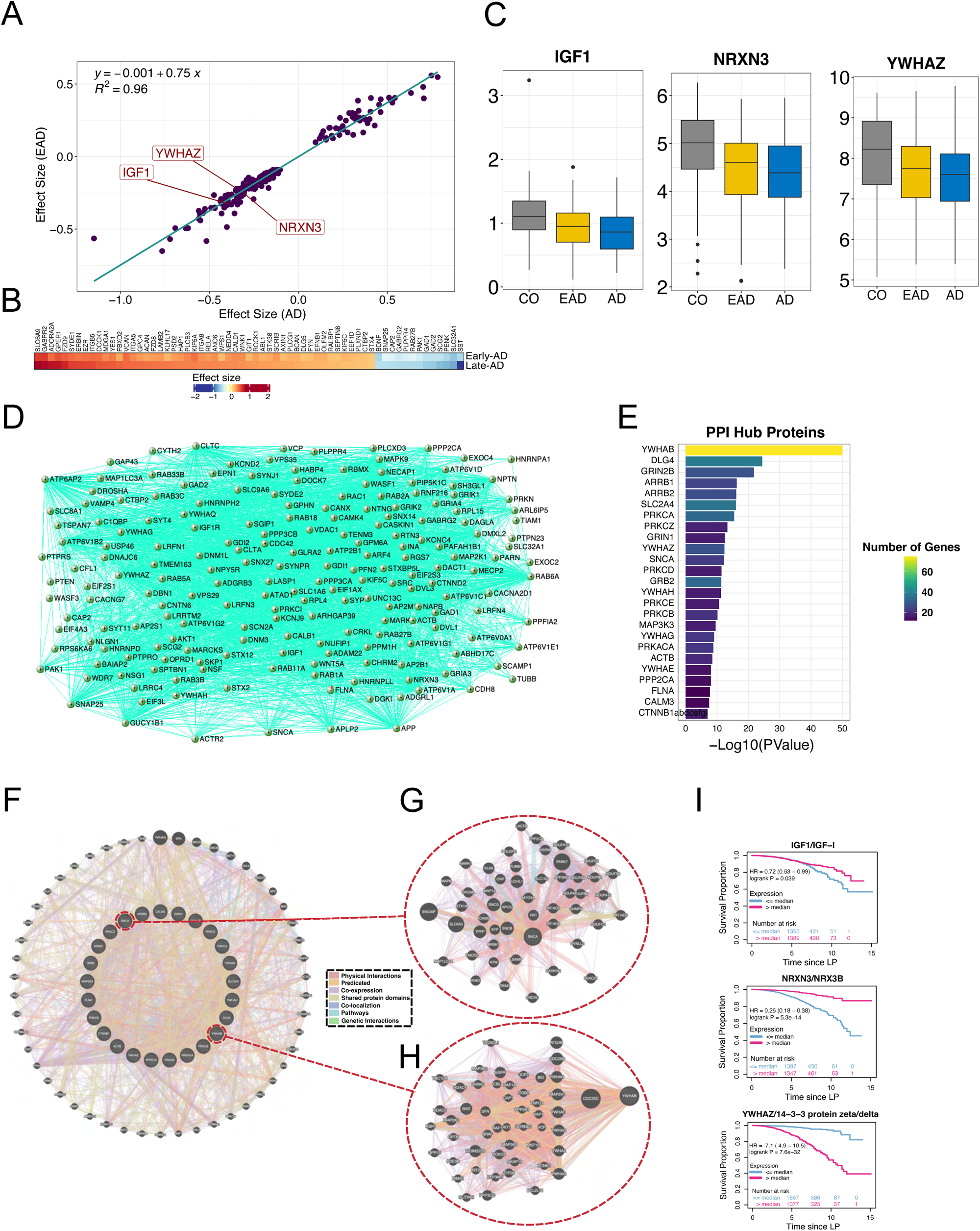
Synaptic dysregulation at multiple stages of AD identified CSF synaptic biomarkers for the molecular staging of AD. (A) Scatterplot showing the correlation of effect size between early-AD (EAD) and late-AD (AD) for common dysregulated synaptic genes in Knight-C4 and RSOSMAP. (B) Heatmap showing the effect size similarity of the most significant synaptic genes detected in EAD and AD. (C) Boxplots showing a consistent expression (transcriptomic) pattern between control, early-AD, and late-AD for genes *IGF1*, *NRXN3*, and *YWHAZ*. (D) The gene interaction network for the turquoise module shows high interactions between dysregulated synaptic genes such as *SNAP25*, *SNCA*, and *APP*. (E) Barplot showing the top 25 hub proteins extracted from protein-protein interaction (PPI) hub proteins pathways. (F) Gene interaction network showing the relatedness of the 25 dysregulated hub proteins. The plot demonstrates multiple strong interactions, including physical and genetic interactions among the query genes (25 hub proteins) and other genes. (G) *YWHAB* sub-network shows the genes highly connected with YWHAB, some of which are examples of either synaptic or other AD-related genes. (H) Same as “g” but for *SNCA* sub-network. (**i**) Kaplan-Meier plots showing the association between *IGF1*, *NRXN3*, and *YWHAZ* genes and an increased risk of dementia progression using CSF proteomics data.

Next, we sought to determine whether some of these genes are potential CSF biomarkers for staging AD pathology and monitoring disease progression. To explore this hypothesis, we performed survival analyses, including individuals with CDR change from 0 to 0.5, and using CSF proteomic (Somascan) data from participants from the Knight ADRC. Our results showed that 11 of 13 (84%) synaptic genes were associated with an increased risk of dementia progression, including *IGF1*, *NRXN3*, and *YWHAZ* (**Fig 9I**). Several studies have shown the involvement of *IGF1* in regulating signaling pathways altered in AD and neurodegeneration [115–119] and its potential as an early CSF/plasma biomarker of disease onset [120]. However, the association between *IGF1* and the increased risk of dementia progression in different stages of AD has not been studied. Likewise, the low expression of *NRXN3* is associated with an increased AD risk [121,122]. The *YWHAZ/ 14−3−3* protein, present in CSF patients, has also been shown to be involved in the rapid progression of dementia [123] and identified as a promising therapeutic target for AD intervention [124]. Results show that the multimodal clusters identified by the cross-omics approach in post-mortem tissue could identify molecular CSF biomarkers for monitoring AD progression in live patients.

## Discussion

In the current study, we leveraged machine learning approaches to integrate high-throughput multi-omics data from controls and AD cases in multiple cohorts and brain regions. Our results highlight the utility of integrating multi-omics data and indicate that cross-omics signatures can better capture differences and discriminate molecular variations in complex and heterogenous diseases such as AD. We found a distinct molecular profile associated with worse cognitive function, earlier age at onset/death, and dysregulated neuronal/synaptic pathways in the parietal cortex. Our results suggest that this multi-omics molecular profiling captures novel molecular events not fully captured by β-amyloid or tau staging as previously reported [125,126]. Similar to previous studies^8^, we did not find a significant association of *APOE* ε4 or PRS with any molecular profile, suggesting that these molecular profiles are not driven by genetic factors associated with AD risk. Through a rigorous and stringent replication using multiple cohorts (MSBB, ROSMAP), we also determined that this molecular profile replicates in additional cortical regions (parietal cortex, DLPFC, PHG) where it associates with tau pathology. Further, we demonstrated that this molecular profile is associated with poor clinical and cognitive function in these studies, offering new insights into the genes/proteins/metabolites associated with cognition decline and pathways associated with worse prognosis.

Notably, we observed that these donors with poor cognitive function, earlier age of onset, and younger age at death showed significantly increased cortical cellular proportions of astrocytes and reduced proportions of neurons. This subset of AD-afflicted participants is significantly enriched in dysregulated synapse-related genes and pathways, which may reflect greater synaptic losses, including dopaminergic synapse and glutamatergic pathways. Although the involvement of dopamine in AD is still debatable, several studies have shown an association between AD pathologies (e.g., Aβ deposition) and dopaminergic dysfunction [127–135]. Similarly, increasing evidence has demonstrated an association between glutamate alterations and AD pathology [136–139]. In addition, transcriptomic and proteomic DE analyses identified several genes associated with AD, including *SNAP25*, *GFAP* and *CLU,* that are more noticeably dysregulated in the distinct molecular profiles. We showed that lower levels of CSF *CLU* are associated with an increased risk of dementia progression in an extended cohort from the Knight ADRC. Moreover, metabolic differential abundance analyses identified significant metabolites associated with biologically relevant AD pathways, most of which are related to major neurotransmitter pathways. These brains also exhibited a metabolic profile associated with disease duration, supporting its implication in the later stages of AD. These novel molecular findings can be leveraged to identify new biomarkers and potential therapeutic targets – not necessarily related to amyloid and tau – that might be effective even in the later stages of the disease.

By integrating single-nuclei RNA-seq, we extracted additional insights supporting cell-type distinct patterns implicated in molecular AD profiles. For instance, we determined that *CX3CR1*, a homeostatic microglia gene, is significantly downregulated in Knight-C4. The lower expression of *CX3CR1* may indicate fewer homeostatic microglia, leading to neuronal damage and loss. The association between homeostatic microglia function and the degree of neuronal cell loss is linked to AD [140]. More recently, the loss of *CX3CR1* was associated with microglial dysfunction illustrated by dampened TGFβ-signaling, increased oxidative stress responses, and dysregulated pro-inflammatory activation^136136^. However, we could not replicate a significant overexpression of microglia-activated genes in additional cortical regions.

We identified *SNCA/α*Syn as one of the top molecules for discriminating AD profiles. We demonstrated that the transcriptomic/proteomic levels of *SNCA* were under-expressed in a subset of AD cases with more advanced tau pathology and worse cognition and replicated this finding in several cohorts and brain regions. Despite the substantial efforts in studying the association between this protein and AD pathology, the mechanisms by which *αSyn* contributes to the pathogenesis and progression of AD remain elusive. Several regulatory mechanisms that contribute to *αSyn* levels have been proposed. For instance, previous studies based on primary neuronal cultures [142], mouse models [143], and live cell imaging [144] have shown that neuronal or synaptic activities regulate the aggregation of *αSyn*. The downregulation of *SNCA/α*Syn in a subset of AD cases with unfavorable outcomes suggests that *SNCA* might be a marker of neuron/synapse losses that can be used for identifying AD stages.

We performed AD staging analyses to study the progression of AD and its effect on the synaptic activities using dysregulated synaptic genes in the Knight-C4 and ROSMAP-C1 profiles. Our data suggest that synaptic dysfunction occurs at the early stages of the disease, but it is more pronounced as AD progresses. Survival analyses from CSF proteomics from the Knight ADRC identified several synaptic genes associated with an increased risk of dementia, such as *IGF1*, *NRXN3*, and *YWHAZ*. Those proteins can be used as potential CSF biomarkers to monitor disease progression.

Although this study provides new insights for understanding the molecular heterogeneity in AD brains, our study has several limitations, highlighting critical future research directions. A limited number of brain metabolomic studies were available, and these were largely underpowered. In addition, the multiple omics data from different brain regions and different cohorts were generated at different times on different samples using different platforms. Thus, future studies that consistently apply the same platforms on multiple brain regions and cohorts can provide novel and deeper findings. We also had to integrate omics data using different platforms due to the need for more harmonization in the methods used by different groups. Independent replication is needed to validate and better understand the role of key drivers of these profiles using disease models (e.g., iPSC or mouse models), including the role of *SNCA* in AD and its association with worsening cognitive function in AD. Longitudinal data from CSF/plasma from large cohorts could provide novel insights into the role of dysregulated genes/proteins/metabolites in AD progression. Additional multi-omics studies exploring multiple brain regions, particularly resilient areas, would be exceptionally useful to determine the temporal progression of protein changes throughout AD progression. We only used one tool (iClusterBayes) to integrate large multi-omics data; however, developing novel and more statistically robust tools is needed. We plan to continue integrating additional cohorts and omics data (e.g. epigenomics) as they become available. Another promising area for future exploration is integrating cross-omics data with drug repositioning datasets that may lead to identifying novel therapeutic potentials for AD, bringing AD closer to precision medicine.

In summary, we have shown the capability of cross-omics approaches and its superiority over single-omic analyses as a powerful resource that provides molecular insight into the AD pathogenesis and identifies possible biomarkers for early synaptic dysfunction, cognition, and AD staging that may eventually enable precision medicine.

## Materials and Methods

### Study Cohorts

#### Discovery (Knight ADRC)

Frozen postmortem parietal lobe tissue samples from Knight Alzheimer Disease Research Center (Knight ADRC) participants, collected with informed consent for research use, were provided by the Knight ADRC Neuropathology Core. The institutional review board of Washington University in St. Louis approved analysis of these samples. Only sporadic AD participants and controls with transcriptomics, proteomics, and metabolomics data available were included in this study, resulting in 255 sporadic AD and 23 controls.

##### RNA-seq

The data generation for the RNA-seq dataset has been previously described [10–12]. Briefly, total RNA was extracted from frozen parietal cortex tissue using a Tissue Lyser LT and purified using RNeasy Mini Kits (Qiagen). The Nanodrop 8000 (Thermo Scientific) and TapeStation 4200 (Agilent Technologies) were used to perform quality control of the RNA’s concentration and purity, and degradation. The RNA integrity number (RIN) was calculated using an RNA 6000 Pico assay on a Bioanalyzer 2100 and TapeStation 4200 (Agilent Technologies). The software determines the RINe on the Bioanalyzer and TapeStation taking into account the entire electrophoretic trace of the RNA, including the presence or absence of degradation products. The DV200 value is defined as the percentage of nucleotides greater than 200nt. All cDNA libraries were prepared using a TruSeq Stranded Total RNA Sample Prep with Ribo-Zero Gold kit (Illumina) and sequenced on an Illumina HiSeq 4000 using 2 x 151 paired-end reads at the McDonnell Genome Institute at Washington University in St. Louis. The average number of reads per sample was 43,195,080.

##### Proteomics

Proteomic data from the Knight ADRC were generated on the SomaLogic platform (SomaLogic Operating Co., Inc., Boulder, CO) from frozen parietal cortical tissue samples (∼500mg) and CSF samples. The SomaLogic platform is a multiplexed, aptamer-based protein quantification platform [145]. The platform measured 1,305 aptamers in total. Full details of the proteomic data generation have been described previously [13].

##### Metabolomics

Metabolomic data from the Knight ADRC were generated on the Metabolon Precision Metabolomics^TM^ UPLC-MS/MS platform (Metabolon, Inc., Morrisville, USA) from 50mg frozen parietal cortical tissue samples. The platform measured 880 metabolites from nine “super pathways”: amino acids, carbohydrates, cofactors and vitamins, energy, lipids, nucleotides, peptides, xenobiotics, and partially characterized molecules. Metabolon provided structural identity annotations for 815 metabolites; the remaining 65 were excluded from the final dataset. Full details of the metabolomics data generation have been described previously [14].

##### Single-nuclei (snRNA-seq)

The data generation of snRNA-seq has been previously described in [146,147]. Briefly, 74 frozen parietal tissues were processed according to the ‘Nuclei extraction and library preparation’ protocol described in [146]. The tissue was homogenized in this protocol, and the nuclei were isolated using a density gradient. The nuclei were then sequenced using the 10X Chromium single-cell Reagent Kit v3, with 10,000 cells per sample and 50,000 reads per cell for each of the 74 samples.

#### Replication (MSBB, BM36)

##### RNA-seq

The RNA-seq raw data from The Mount Sinai Brain Bank (MSBB) study was downloaded from the Synapse portal (syn3157743). It is publicly available as part of the Accelerating Medicines Partnership for Alzheimer’s Disease (AMP-AD). In brief, these data were generated and sequenced from RNA extracted from four postmortem brain regions, including frontal pole (BM10), superior temporal gyrus (BM22), parahippocampal gyrus (BM36), and inferior frontal gyrus tissue (BM44) from 313 subjects. RNA-seq libraries were prepared using the TruSeq RNA Sample Preparation Kit v2 (Illumina, San Diego, CA). The rRNAs were depleted using the Ribo-Zero rRNA Removal Kit (human/mouse/rat) (Illumina). Single-end non-standard reads of 101bp were generated by Illumina HiSeq 2500 (Illumina) [34]. The average number of reads per sample was 32,201,200. RNA-seq data from the region parahippocampal gyrus (BM36) were selected for the cross-omics integration approach because of the availability of proteomics data. Data from the remaining regions were used to explore the identified profiles’ expression profiles to determine whether these are consistent across cortical brain regions.

##### Proteomics (TMT)

The proteomics TMT data were obtained from the AMP-AD Synapse portal (syn25006650) processed by Johnson *et al.*, 2022 [148]. The raw proteomic TMT data is publicly available at AMP-AD and can be downloaded from the Synapse portal (syn21347564). In short, before TMT labeling, the 198 samples were randomized by co-variates (protein quality, sample concentration, diagnosis, age, and sex) into 20 batches (10 cases per batch). The digested peptides were resuspended in 50 mM HEPES (pH 8.5) and labeled with the TMT 11-plex kit (Thermo Fisher) according to the manufacturer’s protocol. In each batch, GIS samples were labeled using TMT channel 126. All 11 channels were mixed equally and desalted with a 100 mg C18 Sep-Pak column (Waters) for the subsequent fractionation [148,149].

#### Replication (ROSMAP, DLPFC)

##### RNA-seq

The RNA-seq raw data from The Religious Orders Study and Memory and Aging Project (ROSMAP) Study were downloaded from the Synapse portal (syn17008934) available from AMP-AD. Briefly, RNA-seq samples were extracted using Qiagen’s miRNeasy mini kit (cat. no. 217004) and the RNase-free DNase Set (cat. no. 79254) and quantified by Nanodrop. Agilent Bioanalyzer evaluated quality. Library preparation was performed by poly-A selection followed by first strand-specific cDNA synthesis, then dUTP for second strand-specific cDNA synthesis, and fragmentation and Illumina adapter ligation for library construction. Illumina HiSeq generated paired-end read sequences with a length of 101bp. Fifty-seven samples were submitted later to the platform and run on an updated protocol (batch 2 protocol). For consistency, we removed those samples from this cohort. The average number of reads per sample was 47,434,600. Only No Cognitive Impairment (NCI) and AD with NO other cause of cognitive impairment cases were used in this study. All cases with “unknown” BraakTau scores were discarded. Plaque density and Neurofibrillary tangle burden (PHF tau tangles) are a summary of AD pathology derived from three AD pathologies: neuritic plaques, diffuse plaques, and neurofibrillary tangles based on five regions: mid-frontal cortex, mid-temporal cortex, inferior parietal cortex, entorhinal cortex, and hippocampus. Tangles are identified by molecularly specific immunohistochemistry based on the mean of eight regions: tangles_hip (hippocampus), tangles_ec (entorhinal cortex), tangles_mf (mid frontal cortex), tangles_it (inferior temporal), tangles_ag (angular gyrus), tangles_calc (calcarine cortex), tangles_cg (anterior cingulate cortex), angles_sf (superior frontal cortex).

##### Metabolomics

Data from the Religious Orders Study and Memory and Aging Project (ROSMAP) were generated by the Duke Metabolomics and Proteomics Shared Resource, a member of the ADMC, using protocols published previously for blood samples [150–152]. A custom protocol developed for the brain samples can be found on Synapse at syn10235609. The dorsolateral prefrontal cortex (DLPFC) metabolomic data from the ROSMAP studies quantified on the Metabolon Precision Metabolomics platform and preprocessed by the ADMC as described in [153] were downloaded from Synapse in July 2021 (syn25878459). The platform measured a total of 1,055 metabolites in the ROSMAP dataset.

### Quantification and statistical analysis

#### RNA-seq QC, alignment, and gene expression quantification

All RNA-seq datasets from the discovery and replication were processed and aligned using our in-house RNA-seq pipeline (https://github.com/HarariLab/RNA-seq-Pipeline). Genome reference and gene models were selected similar to the TOPmed pipeline (https://github.com/broadinstitute/gtex-pipeline/blob/master/TOPMed_RNAseq_pipeline.md). Reference genome GRCh38 and GENCODE 33 annotation, including the addition of ERCC spike-in annotations, were used. We excluded ALT, HLA, and Decoy contigs from the reference genome due to insufficient RNA-seq tools that properly handle these regions. Before alignment, the quality of raw read sequences for all libraries was assessed using FastQC (v0.11.9) [154]. All raw read sequences were aligned to the human reference genome (GRCh38) using STAR (v.2.7.1a) [155]. The alignment quality was evaluated using sequencing metrics such as reads distribution, ribosomal content, or alignment quality provided by STAR using Picard tools (v.2.8.2) [156]. All samples that failed to pass the QC or were outliers (Knight ADRC=9, MSBB=18, ROSMAP=6) were removed from the downstream analyses. Raw read counts for transcripts and genes were generated using STAR and computed transcript/gene expression levels as normalized in FPKM (Fragments Per Kilobase of transcript per Million mapped reads) format.

#### Metabolomics QC and Quantification

Metabolomics data from Knight ADRC and ROSMAP were processed similarly. Full details of the QC process have been previously described [157]. Briefly, 188 metabolites with readings missing in over 20% of donors were excluded, readings were log10-transformed, means per metabolite were adjusted to zero, and outlier readings (outside 1.5xIQR) were excluded. Missing metabolite values in < 20% of donors were replaced by their mean value. No samples were removed due to the missingness rate. Principal component analysis was performed with the FactoMineR R package [158] to identify outlier samples; four outlier samples were excluded. The final dataset consisted of 627 metabolites. A similar procedure was used to clean the ROSMAP metabolomic dataset. Metabolites without assigned structural identities and metabolites with greater than 20% missing readings were excluded, readings were log10-transformed, the mean of each metabolite’s distribution was adjusted to zero, and outlier readings were removed. Two samples missing greater than 20% of readings were excluded. The final dataset consisted of 595 metabolites.

#### Proteomics QC and Quantification

Proteomic data for brain and CSF samples from the Knight ADRC were first filtered based on a limit of detection (LOD). Samples with greater than 15% outlier analytes (analyte readings below the LOD) were excluded. Analytes were then filtered based on scale factor difference, meaning the absolute value of the maximum difference between the calibration scale factor per aptamer and the median for each plate run. Analytes were excluded if their scale factor difference exceeded 0.5. Based on log10-transformation of protein readings, samples exceeding 1.5-fold of the interquartile range (IQR) per analyte were considered outliers. Analytes with greater than 15% outlier samples were excluded. Similarly, samples that were outliers for greater than 15% of analytes were excluded. Full details of the proteomic data QC have been described previously [13]. Missing analyte readings and removed outlier readings were imputed using the impute.knn function from the impute R package (v1.56.0) [159]. A plate-wise batch correction was performed using the ComBat function from the sva R package (v3.30.1) [160]. The final brain dataset comprised 1,092 analytes, and the final CSF dataset consisted of 713 analytes. Before using these data, analytes with missing expressions in > 20% of donors were excluded, and missing expressions of those in < 20% were replaced with their mean value. The processed data for TMT proteomics from MSBB BM36 was obtained from AMP-AD (syn25006647) and processed by Johnson *et al.*,2022 [148]. In brief, 760 raw files generated from 19 TMT 11-plexes were analyzed using the Proteome Discoverer suite (version 2.3, Thermo Fisher Scientific). The UniProtKB human proteome database containing Swiss-Prot and TrEMBL human reference protein sequences was used to search mass spectra. Peptide spectral matches and peptides were filtered to a false discovery rate (FDR) of less than 1% using Percolator software. After spectral assignment, peptides were assembled into proteins and filtered based on the combined probabilities of their constituent peptides to a final FDR of 1% [148]. Unique and razor (that is, parsimonious) peptides were considered for quantification. TMT reporter abundance was transformed into a ratio followed by log_2_ transformation. Quality control analysis was performed, and outliers were removed. Similar to the Knight ADRC, analytes with missing expressions in > 20% of donors were excluded, and missing expressions of those in < 20% were replaced with their mean value.

#### Single-nuclei (snRNA-seq) QC, alignment, and gene expression quantification

Full details of the data process of snRNA-seq data have been previously described in [146,147]. In short, CellRanger (v2.1.1 10XGenomics) [161] software was employed to align the sequences and quantify gene expression. Read sequences were aligned to a custom pre-mRNA reference constructed from genome assembly GRCh38 and generated as described by 10X Genomics technical manual. Filtering and QC analyses were performed individually using each subject’s Seurat package (v2.20 and 2.30). All nuclei with high mitochondria gene expression were removed, and only genes expressed in > 3 cells were kept for downstream analysis. Cells with < 1800 or > 8000 genes expressed in them were excluded. The data were normalized using the *LogNormalize* function that normalizes the gene expression measurements for each cell by the total expression, scales by a factor equal to the median counts of all genes, and log-transforms the expression.

### Clustering of AD participants

To cluster AD participants, iClusterBayes (Integrative clustering of multiple genomic data types) [33] available from iClusterPlus R package (v1.22.0) [162] was employed. For each integration process, shared samples across omics datasets were extracted, and the top 2000 most variant features were selected. iClusterBayes was performed by fitting multiple Bayesian models using different clusters ranging from 1 to 10. We ran 22,000 Markov Chain Monte Carlo (MCMC) iterations for each, of which the first 12,000 were discarded as burn-in. The prior probability for the indicator variable gamma of each data set was set to 0.1, whereas the posterior probability cutoff was set at 0.5. The standard deviation of the random walk proposal for the latent variable was set to 0.5. The remaining parameters were left with the default values. We used the Bayesian information criteria (BIC) and deviance ratio to evaluate clustering solutions and maximizing the compactness of the clusters (intra-cluster similarity).

### Cell proportion analysis

CellMix R package (v1.6.2) [163] employs multiple digital deconvolution methods and was used to estimate cellular population structure from gene expression data (TPM) generated by the Salmon quantification tool. The machine learning model and the gene panel (genes marker expressed highly in specific cell types) were previously described in [12]. Four cell types (astrocytes, microglia, neurons, and oligodendrocytes) were tested using two algorithms, meanProfile [163] and ssNMF (semi-supervised nonnegative matrix factorization) [164]. The significant difference in cell proportion between clusters was computed using a generalized linear model (GLM) test and corrected by sex and age of death (AOD).

### Differential expression analysis of AD clusters

RNA-seq: differential expression (DE) analyses were performed to compare participants in each cluster to the control participants and to other AD clusters using the DESeq2 R package (v.1.22.2) [165]. Our models were adjusted for sex, age at death, and the percentage of astrocytes and neurons. Lowly expressed genes were removed to enhance our confidence in the differentially expressed genes we discovered. Only genes with expression > 0.5 CPM (count per million) in at least 25% of samples in either group being compared were retained for downstream analyses. The false discovery rate (FDR) was estimated using Benjamini-Hochberg (BH) [166]. All genes with FDR < 0.05 were considered differentially expressed genes.

#### Proteomics

differential expression analyses between clusters were carried out with linear models (R function glm) with age at death and sex as covariates. P-values were adjusted using Benjamini-Hochberg (BH) [166].

#### Metabolomics

differential abundance analyses were also carried out with linear models, with age at death, sex, and post-mortem interval (PMI) as covariates. P-values were adjusted using Benjamini-Hochberg (BH) [166].

#### Single-nuclei

differential expression analyses were performed to compare the overexpression of each gene in each cell type relative to other cell types using glmmTMB R Package (v1.1.3). This approach uses a template builder (glmmTMB) to fit generalized linear mixed models. Our model was corrected by sex covariate. To ensure cell-type specific overexpression, for each cell type, only top genes whose effect size is greater than 90% percentile cutoff were retained for downstream integration analyses.

### Survival analyses

Kaplan-Meier survival analyses and Cox proportional hazards models were used to determine the association of different clusters with survival outcomes and also to determine the association with the risk of dementia progression using survival (v3.2-3) [167] and surviplot (v1.1.1) [168] R packages.

### Significance of overlap

The significance of overlap for all analyses was computed using Fisher’s exact test available from the R stats package (v4.3.0) [169].

### Pathway analysis

All pathway analyses for transcriptomics and proteomics were performed using the EnrichR R package (v3.0) [170] using genes/proteins significantly up-and downregulated between clusters vs. control and between distinct clusters. Two widely used databases, gene ontologies (GOs), related to molecular function, biological process, and cellular components, and KEGG databases, were used. All pathways with an adjusted p-value < 0.05 were considered statistically significant. Similar pathways analyses were performed for single-nuclei (snRNA-seq) using EnrichR using significantly up-and downregulated genes in specific cell types. For metabolomics, pathways analyses were performed using MetaboAnalyst (v5.0) software [171] using the metabolites that were increased- and decreased between clusters and the control and across clusters.

### Replication analysis

Our replication strategy was designed to focus on the molecular profiles identified in the discovery analyses of the Knight ADRC cohort by extracting the significant genes and proteins identified in the Knight ADRC cohort. Using this set of features, iClusterBayes was run on both datasets (MSBB and ROSMAP) using the same parameters used with the discovery cohort.

### NanoString gene expression analysis

The data generation and full details of the QC processes have been previously described [114]. Briefly, A53T αSyn-Tg mice were crossed with human *APOE2*, *APOE3*, or *APOE4* knockin mice or *Apoe* knockout mice to generate A53T mice homozygous for one of the three human *APOE* alleles or completely null for *Apoe*. Sample sizes and genotypes for NanoString gene expression analysis were selected based on αSyn pathology and biochemistry results. RNA was isolated from 12-month-old A53T/EKO (n=10), A53T/E2 (n=2), and A53T/E4 (n=10) mouse midbrain. 793 transcripts were quantified with the NanoString nCounter multiplexed target platform using a customized chip based on the Mouse Neuroinflammation panel (www.nanostring.com). The geometric mean of negative control lanes was subtracted from gene transcript counts, and gene expression data were normalized to the geometric mean of positive control lanes and a list of housekeeping genes [114]. QC analyses, including principal components analyses, were performed using nSolver 4.0 and the Advanced Analysis 2.0 plugin (NanoString). For differential expression analysis, pSyn pathology was expressed as low (< 5% coverage by IHC) or high (> 5% coverage by IHC) for each animal, and APOE genotype and pSyn pathology were selected as predictor covariates. Differentially expressed analyses were performed using linear regression models in R with sex as covariates. P-values were adjusted using Benjamini-Hochberg (BH) [166].

### AD staging analysis

Clinical and pathological data, including Reagan, Cerad, and Braak scores as well as final consensus cognitive diagnosis (cogdx) from the ROSMAP cohort, were used to classify AD cases into early-AD and late-AD as well as other clinical statuses such as presymptomatic, mild cognitive impairment (MCI), and control. Differential expression analyses were performed to compare early and late AD cases to control cases using the DESeq2 R package (v.1.22.2) [165]. Dysregulated synaptic genes from the Knight ADRC profile (Knight-C4) overlapped with the dysregulated synaptic genes from each comparison from ROSMAP, and the difference in effect size of the two groups (early-AD vs. Control and late-AD vs. Control) was examined.

### Co-expression and gene interaction analyses

Co-expression analyses were performed using dysregulated synaptic genes in Knight-C4 using the WGCNA R package (v1.72-1) [172]. Gene interaction analyses were performed using GeneMANIA webserver [173].

## Data availability

Transcriptomics data from the Knight ADRC are available by request from the National Institute on Aging Genetics of Alzheimer’s Disease Data Storage Site (NIAGADS) with accession number ng00083 (https://www.niagads.org/datasets/ng00083). Proteomics data from the Knight ADRC donors are available at the NIAGADS Knight ADRC collection with accession number ng00102 (https://www.niagads.org/datasets/ng00102). Metabolomics data from the Knight ADRC are available at the NIAGADS Knight ADRC collection with accession number NG00113 (https://dss.niagads.org/datasets/ng00113/). The single nucleus data from the Knight ADRC are available at NIAGADS Knight ADRC collection with accession number NG00108 (https://dss.niagads.org/datasets/ng00108/). MSBB transcriptomics and proteomics data are publicly available at Synapse under Synapse IDs syn3157743 (https://www.synapse.org/#!Synapse:syn3157743) and syn25006650 (https://www.synapse.org/#!Synapse:syn25006650) respectively. ROSMAP transcriptomics and metabolomics data are publicly available at Synapse under Synapse IDs syn17008934 (https://www.synapse.org/#!Synapse:syn17008934) and syn25878459 (https://www.synapse.org/#!Synapse:syn25878459) respectively.

## Code availability

The analysis code used for this study will be available at the time of publication.

## Acknowledgments

We thank all the participants, their families, the many involved institutions, and their staff, whose help and participation made this work possible. This work was supported by access to equipment made possible by the Hope Center for Neurological Disorders and the Departments of Neurology and Psychiatry at Washington University School of Medicine.

## ROSMAP (RNA-seq)

Study data were provided by the Rush Alzheimer’s Disease Center, Rush University Medical Center, Chicago. Data collection was supported through funding by NIA grants P30AG10161 (ROS), R01AG15819 (ROSMAP; genomics and RNAseq), R01AG17917 (MAP), R01AG30146, R01AG36042 (5hC methylation, ATACseq), RC2AG036547 (H3K9Ac), R01AG36836 (RNAseq), R01AG48015 (monocyte RNAseq) RF1AG57473 (single nucleus RNAseq), U01AG32984 (genomic and whole exome sequencing), U01AG46152 (ROSMAP AMP-AD, targeted proteomics), U01AG46161(TMT proteomics), U01AG61356 (whole genome sequencing, targeted proteomics, ROSMAP AMP-AD), the Illinois Department of Public Health (ROSMAP), and the Translational Genomics Research Institute (genomic). Additional phenotypic data can be requested at www.radc.rush.edu. Study data were provided through NIA grant 3R01AG046171-02S2 awarded to Rima Kaddurah-Daouk at Duke University, based on specimens provided by the Rush Alzheimer’s Disease Center, Rush University Medical Center, Chicago, where data collection was supported through funding by NIA grants P30AG10161, R01AG15819, R01AG17917, R01AG30146, R01AG36836, U01AG32984, U01AG46152, the Illinois Department of Public Health, and the Translational Genomics Research Institute.

## Alzheimer’s Disease Metabolomics Consortium (ADMC)

The results published here are in whole or partly based on data obtained from the AD Knowledge Portal (https://adknowledgeportal.org). Metabolomics data is provided by the Alzheimer’s Disease Metabolomics Consortium (ADMC) and funded wholly or in part by the following grants and supplements: NIA R01AG046171, RF1AG051550, 3U01AG024904-09S4, RF1AG057452, R01AG059093, RF1AG058942, U01AG061359, U19AG063744, and FNIH: #DAOU16AMPA awarded to Dr. Kaddurah-Daouk at Duke University in partnership with a large number of academic institutions. As such, the investigators within the ADMC, not listed specifically in this publication’s author’s list, provided data along with its pre-processing and prepared it for analysis but did not participate in the analysis or writing of this manuscript. A complete listing of ADMC investigators can be found at: https://sites.duke.edu/adnimetab/team/.

## MSBB

The results published here are in whole or in part based on data obtained from the AD Knowledge Portal (https://adknowledgeportal.org/). These data were generated from postmortem brain tissue collected through the Mount Sinai VA Medical Center Brain Bank. Dr. Eric Schadt from Mount Sinai School of Medicine provided them. Dr. Levey provided proteomics data from Emory University based on postmortem brain tissue collected through the Mount Sinai VA Medical Center Brain Bank provided by Dr. Eric Schadt from Mount Sinai School of Medicine.

## Author’s contributions

A.M.E. conciliated the data, supervised the processing of the RNA-seq, optimized the ML learning approach, performed the analyses, interpreted the data, and wrote the manuscript. A.M.E. and B.C.N. processed, QCed, and analyzed the metabolomics and proteomics data. C.S.T. analyzed and interpreted pathway results. A.M.E. and E.D. processed and QCed all RNA-seq data. L.B. processed and QCed single-nuclei data. A.N. performed NanoString gene expression analyses and prepared all supplementary tables for the manuscript. J.B. and C.S. contributed to the computational analyses and phenotype harmonization. F.F. processed genetic data. K.B., J.B., J.N., J.G., F.W. contributed to data collection, processing, quality control, and cleaning from the Knight ADRC. J.C.M. and R.J.P. contributed samples and/or data. B.A.B., R.J.P., A.A.D., and C.M.K. interpreted the results and provided critical feedback and input to the manuscript. O.H. conceived and designed the study, collected the data, and supervised the project. O.H. and B.A.B. interpreted the results and supervised the drafting of the manuscript. All the authors critically reviewed the manuscript and approved the final version.

## Supporting information

**S1 Fig.**
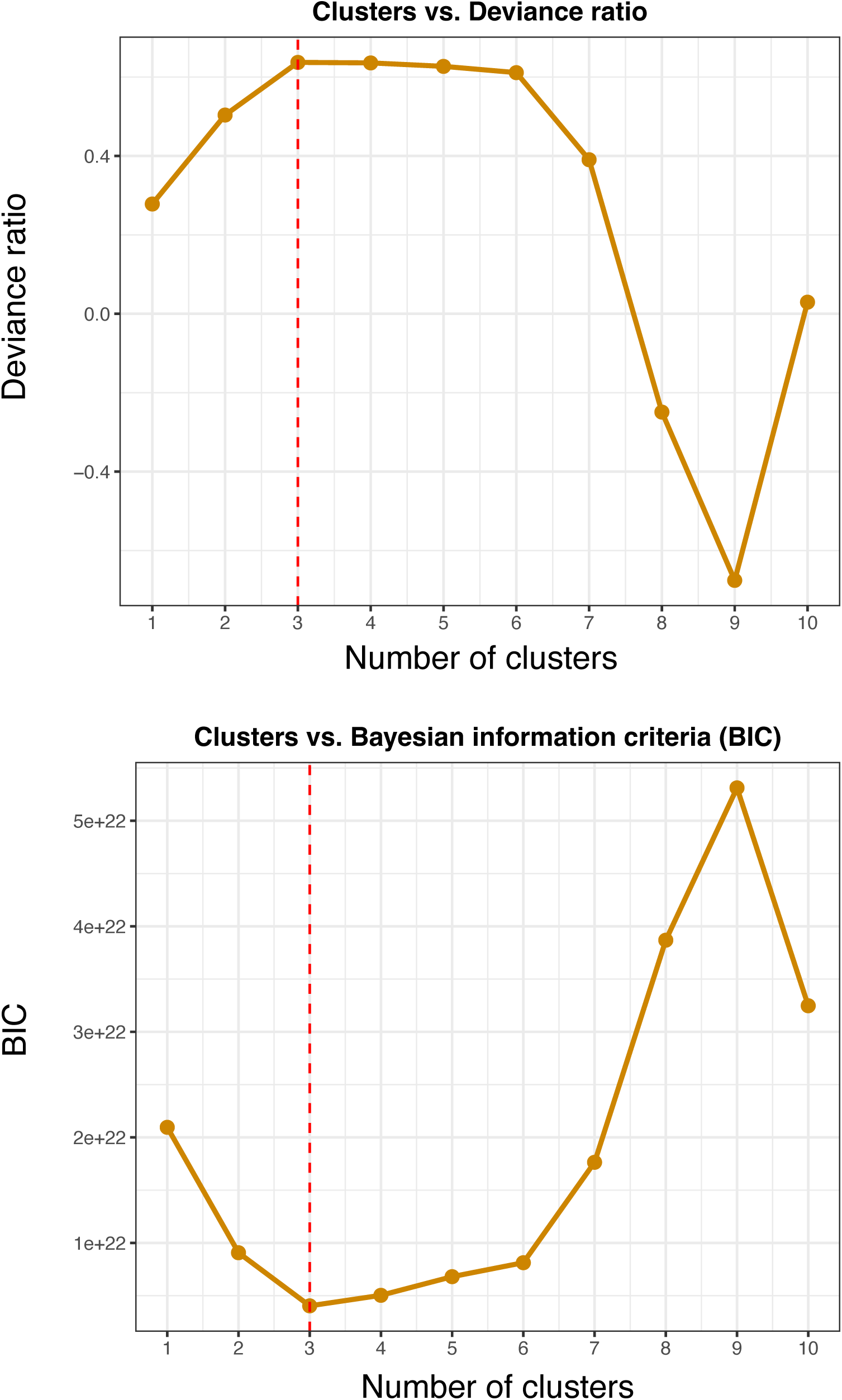
The maximum deviance ratio (left) and the minimum Bayesian information criterion—BIC (right) results across cluster solutions. The optimal solution is observed with cluster number *K=3* dividing our samples into four clusters (*k+1*).

**S2 Fig.**
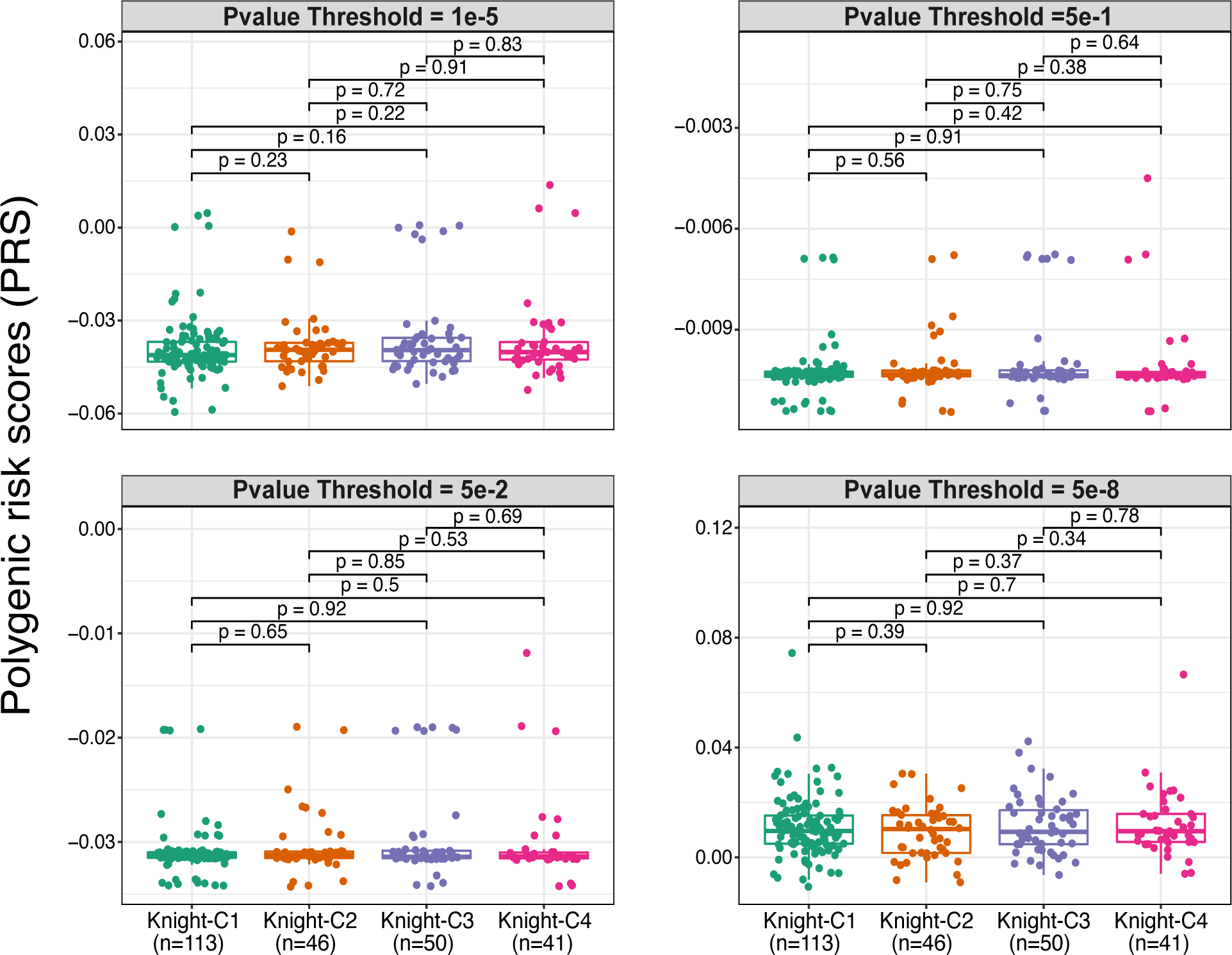
Knight-C4 is not associated with polygenic risk scores (PRS). Boxplots showing the polygenic risk scores (PRS) across four clusters using four different p-value thresholds showing no significant association between Knight-C4 and PRS scores.

**S3 Fig.**
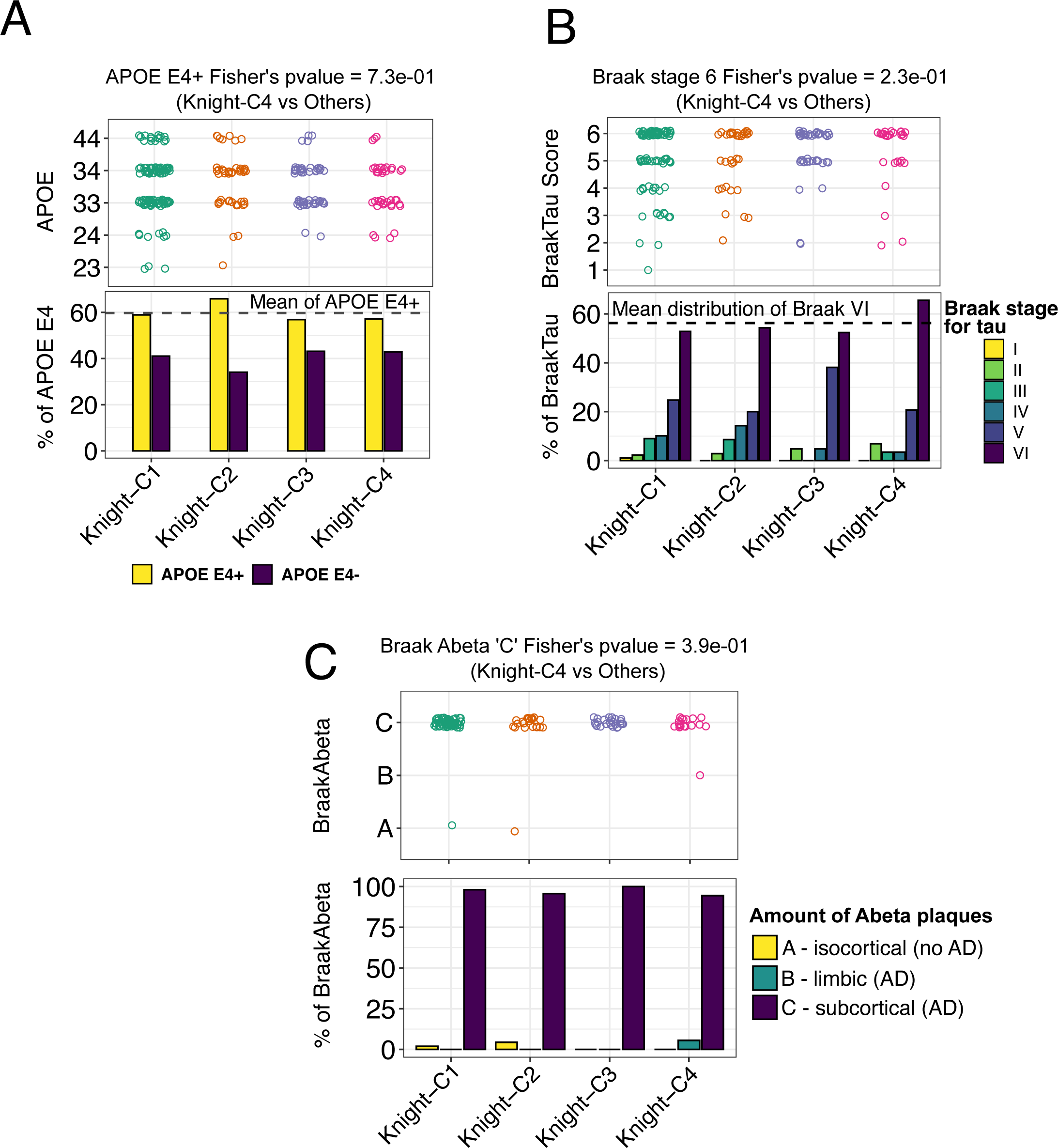
Knight-C4 is not associated with genetic or neuropathological effects. (**a**) The distribution of the APOE ℇ4 allele across four clusters shows no association with Knight-C4. (**b**) The distribution of Braak scores for tau across four clusters shows no significant correlation between Knight-C4 and Braak staging for tau. However, Knight-C4 exhibited more stage VI cases exceeding the mean distribution of Braak stage VI (dashed line) across all clusters. (**d**) The distribution of Braak scores for amyloid-β across four clusters showing no association with Knight-C4.

**S4 Fig.**
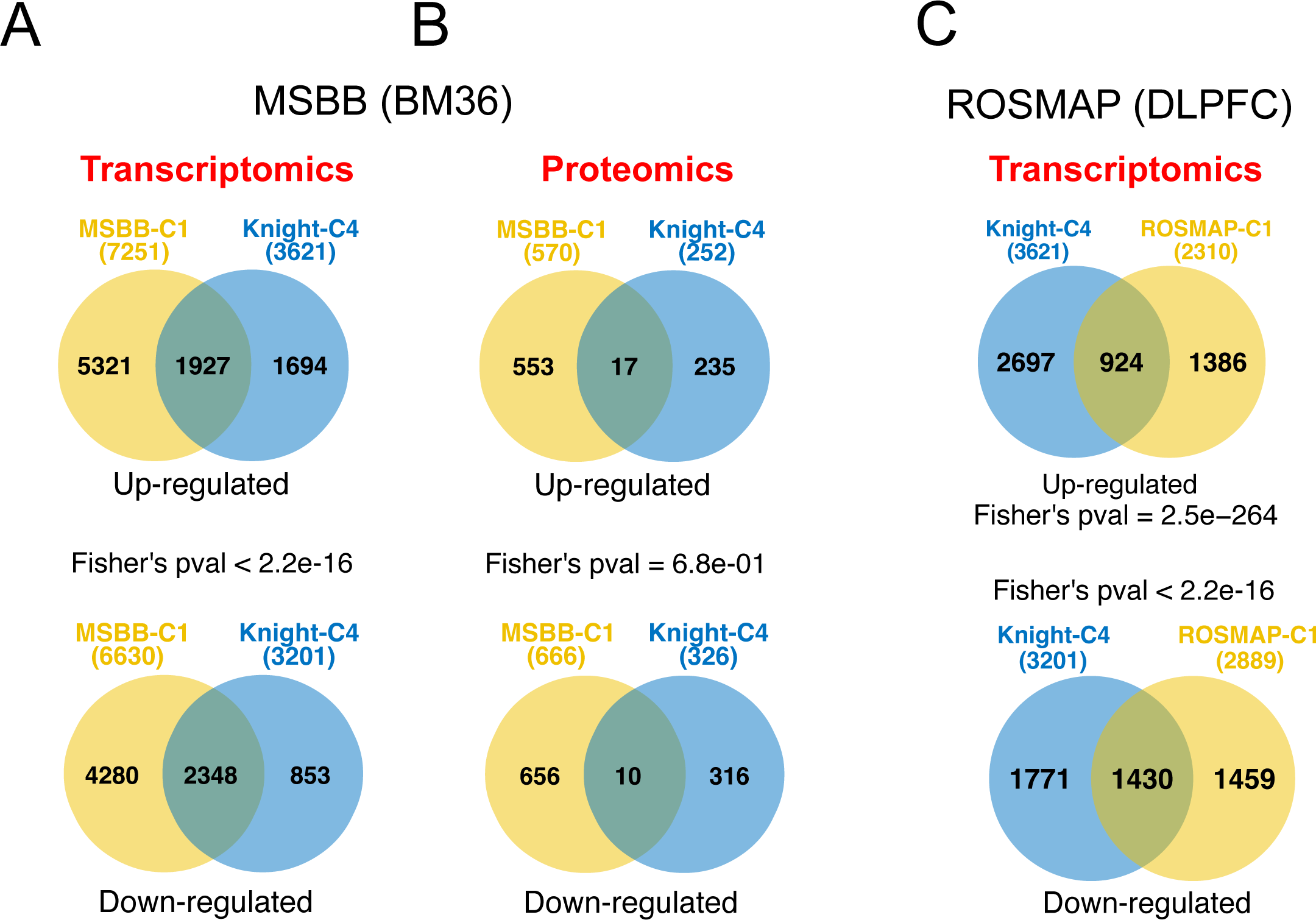
Dysregulated genes are significantly common across cohorts from multiple brain regions. (**a**) Venn diagrams depicting the overlap between significant genes in Knight-C4 and MSBB-C1 cohorts, showing a replication of the molecular signature associated with worse cognitive outcomes. (**b**) Same as “a” but for significant proteins. (**c**) Venn diagrams depicting the overlap between significant genes in Knight-C4 and ROSMAP-C1 cohorts, showing a replication of the molecular signature associated with worse cognitive outcomes.

**S5 Fig.**
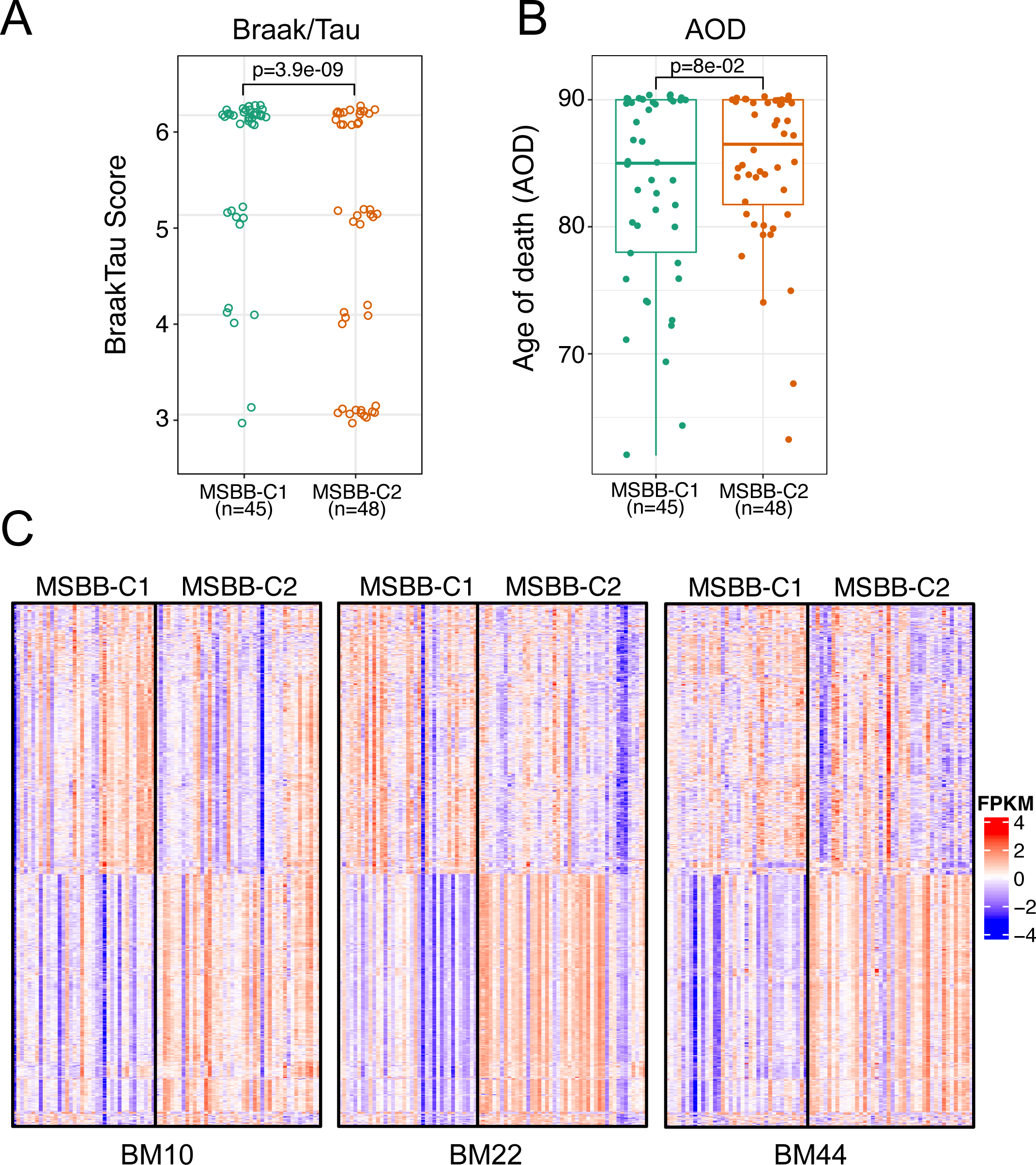
Molecular profiles of MSBB-C1 are associated with high Braak scores for tau and replicated in additional brain regions from the MSBB study. (**a**) Boxplots showing MSBB-C1 are associated with high BraakTau scores. (**b**) Boxplots show a clear pattern of an association with the early age of death for cases in MSBB-C1. (**c**) Heatmaps showing the replication of transcriptomic profiles of MSBB-C1 from the BM36 region in additional regions, including frontal pole (BM10), superior temporal gyrus (BM22), and inferior frontal gyrus (BM44) brain regions from the MSBB study.

**S6 Fig.**
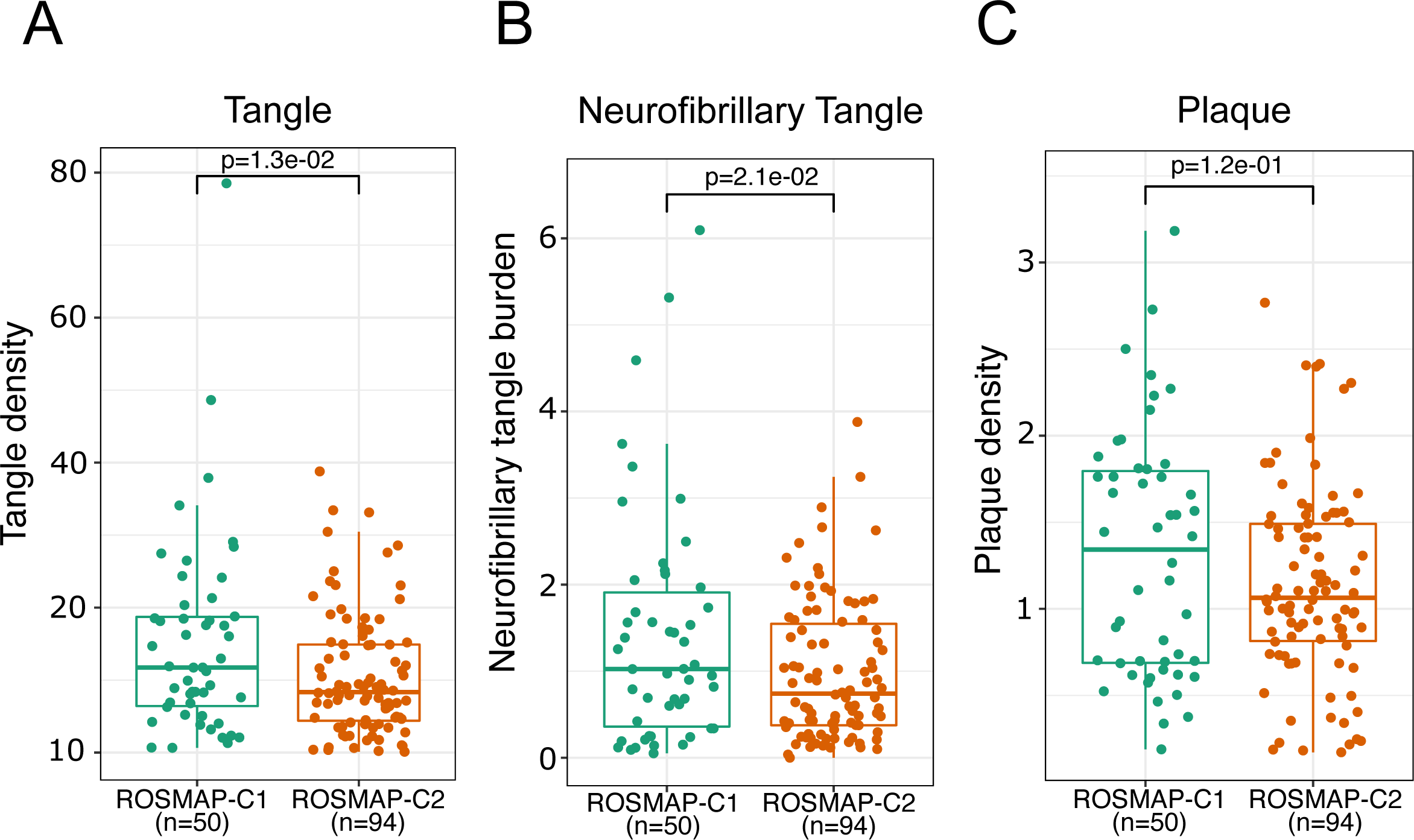
Molecular profiles of ROSMAP-C1 are associated with multiple neuropathological variables. **(a)** Boxplots showing ROSMAP-C1 association with high tangle densities. (**b**) Boxplots showing ROSMAP-C1 association with neurofibrillary tangle burdens. (**c**) Boxplots showing higher plaque densities for AD cases in ROSMAP-C1.

**S7 Fig.**
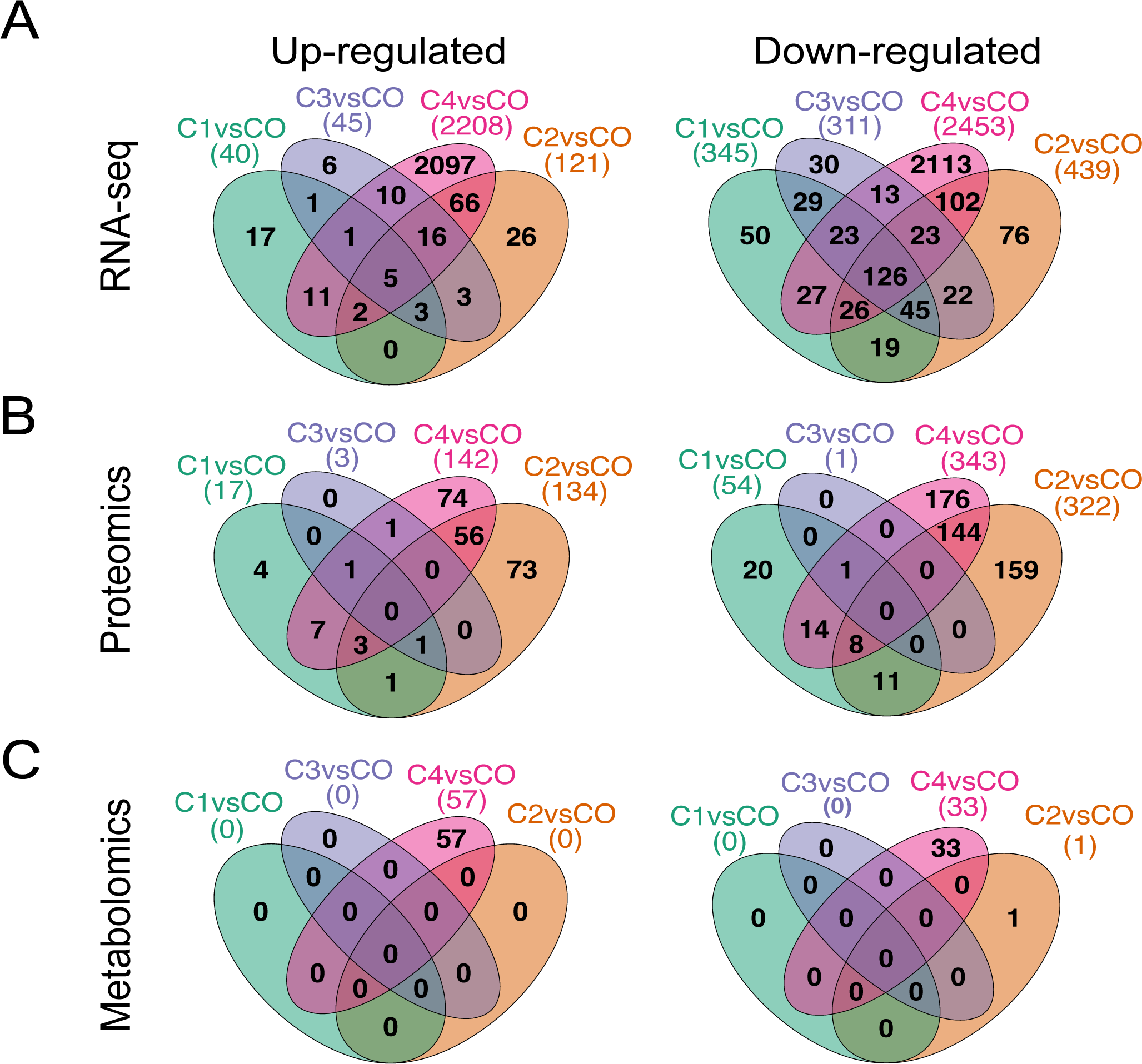
Knight-C4 is associated with more significant hits across multi-omics datasets. (**a**) Venn diagrams show the differentially expressed genes detected in each cluster (cluster vs control). (**b**) Same as panel “a” but for proteomics. (**c**) Same as panel “a” but for metabolomics.

**S8 Fig.**
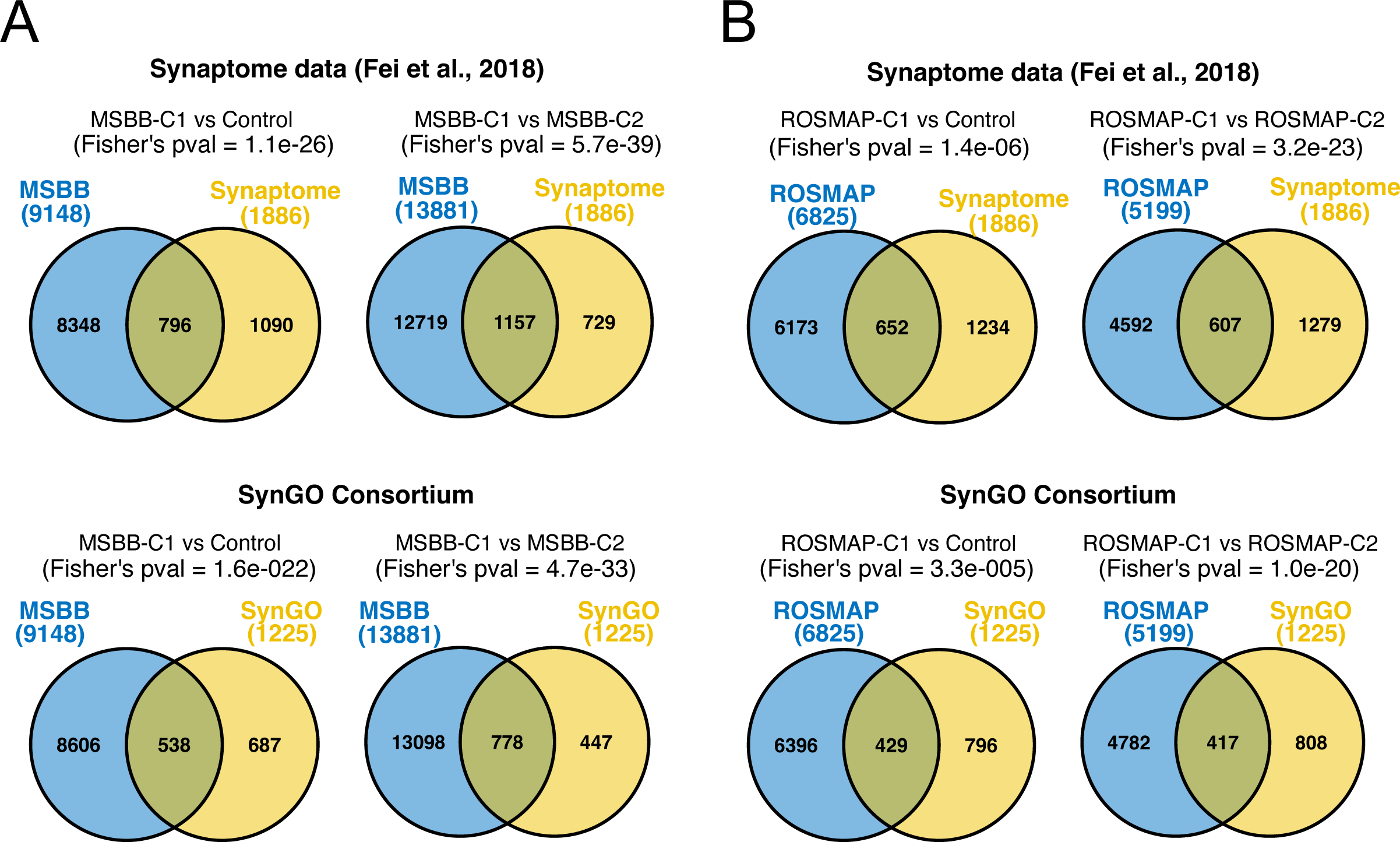
Significant enrichment of dysregulated synaptic genes in Knight-C4 is replicated in additional brain regions. (**a**) Venn Diagrams showing the overlap between significant genes in MSBB-C1 and synaptic genes from Fei *et al.,* 2018 [37] and SynGO [36] datasets. (**b**) Same as “a” but overlaps with significant genes in ROSMAP-C1.

**S9 Fig.**
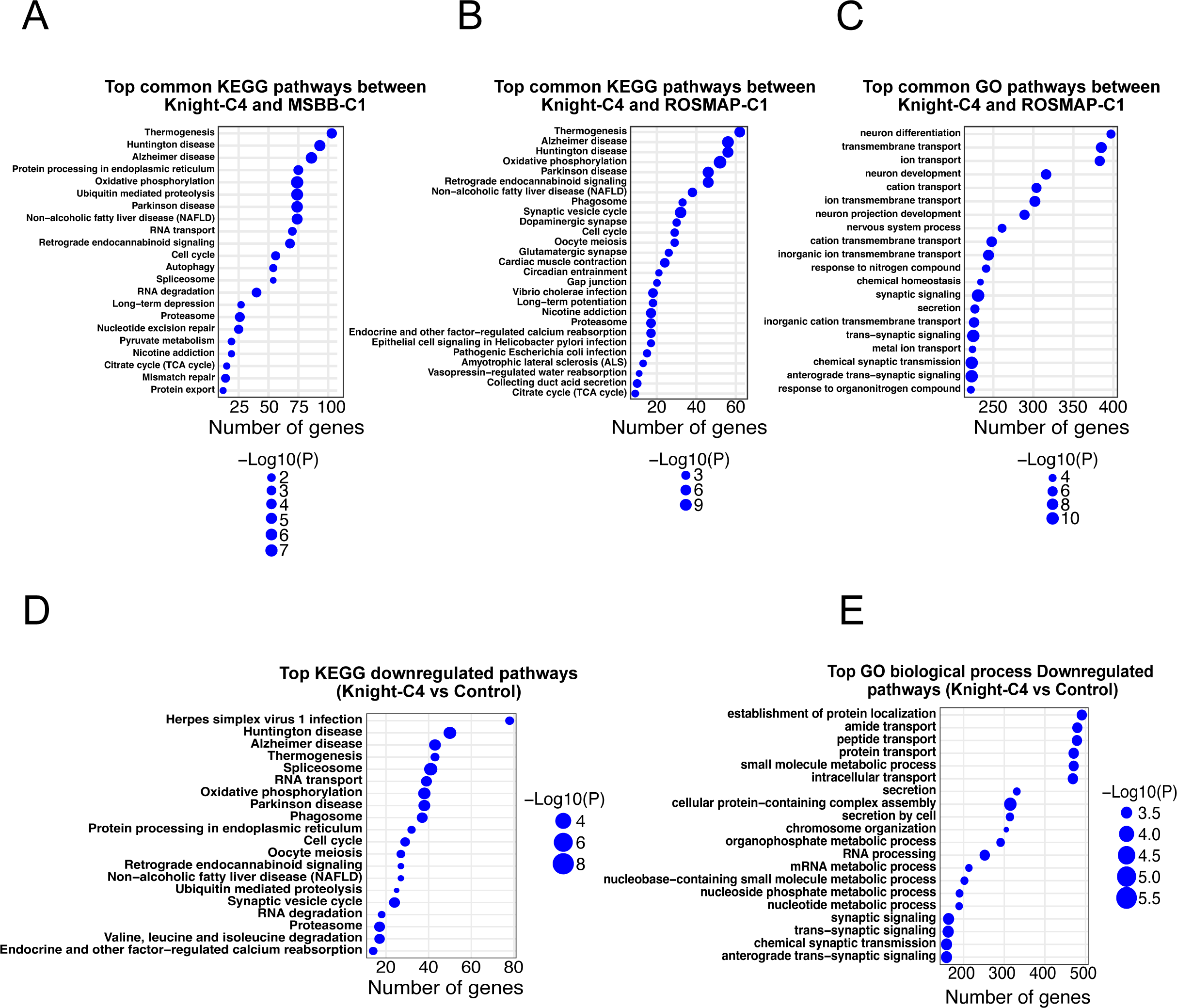
Top significant pathways dysregulated in AD molecular profiles from multiple cortical brain regions and AD cohorts. (**a**). Top common KEGG pathways dysregulated in Knight-C4 and MSBB-C1 (Cluster vs other AD cases). (**b**) Top common KEGG pathways dysregulated in Knight-C4 and ROSMAP-C1 (Cluster vs other AD cases). (**c**) Top common GO biological process pathways dysregulated in Knight-C4 and ROSMAP-C1 (Cluster vs other AD cases). (**d**) Top 20 KEGG pathways enriched in Knight-C4 downregulated genes (Knight-C4 vs control). (**e**) Top 20 GO biological process pathways enriched in Knight-C4 downregulated genes (Knight-C4 vs control).

**S10 Fig.**
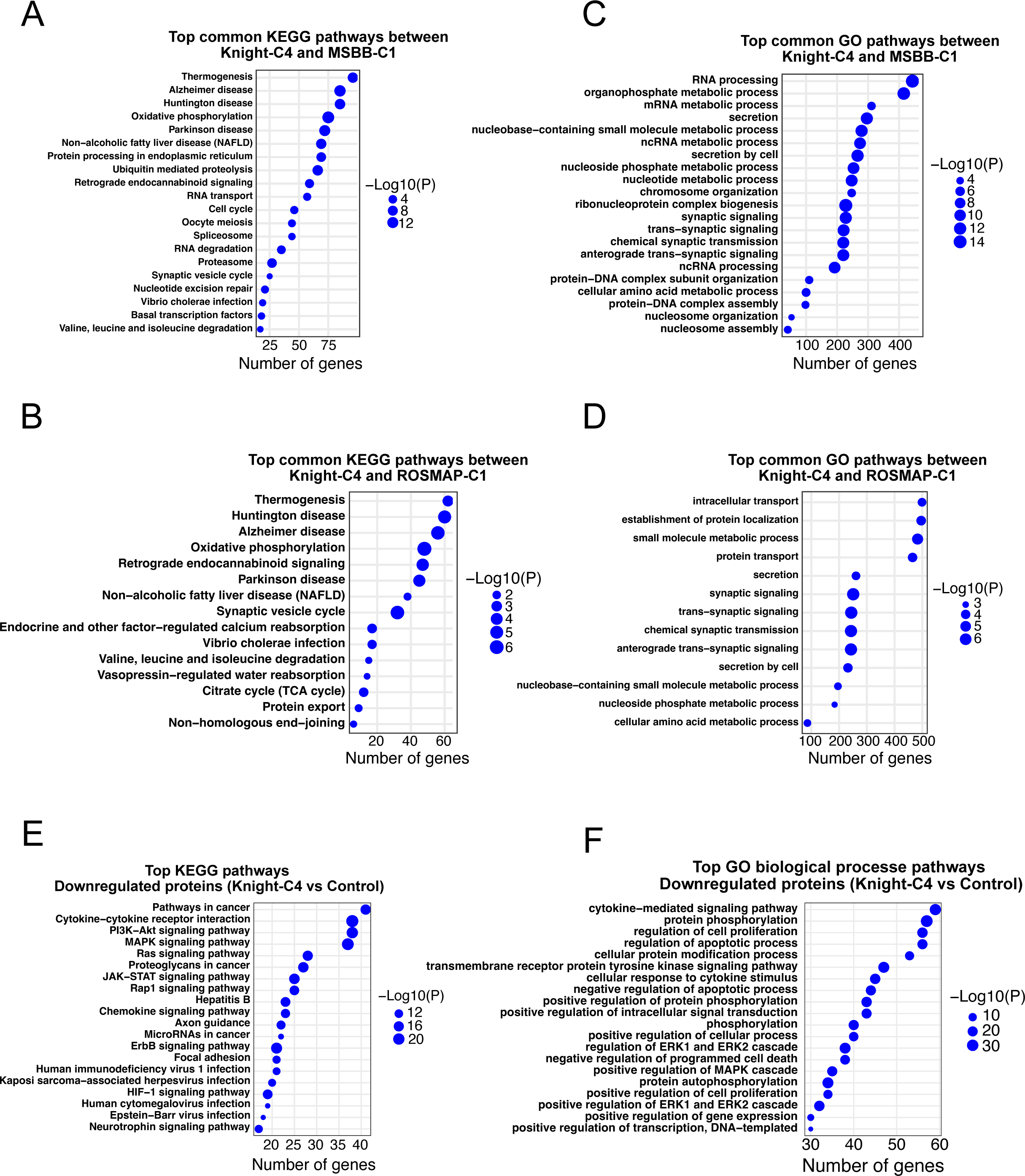
Top significant pathways dysregulated in AD molecular profiles from multiple cortical brain regions and AD cohorts compared to control cases. (**a**) Top common KEGG pathways dysregulated in Knight-C4 and MSBB-C1. (**b**) Top common KEGG pathways dysregulated in Knight-C4 and ROSMAP-C1. (**c**) Top common GO biological process pathways dysregulated in Knight-C4 and MSBB-C1. (**d**) Top common GO biological process pathways dysregulated in Knight-C4 and ROSMAP-C1. (**e**) Top 20 KEGG pathways associated with the downregulation of proteins in Knight-C4. (**f**) Top 20 GO biological process pathways enriched in Knight-C4 downregulated proteins.

**S11 Fig.**
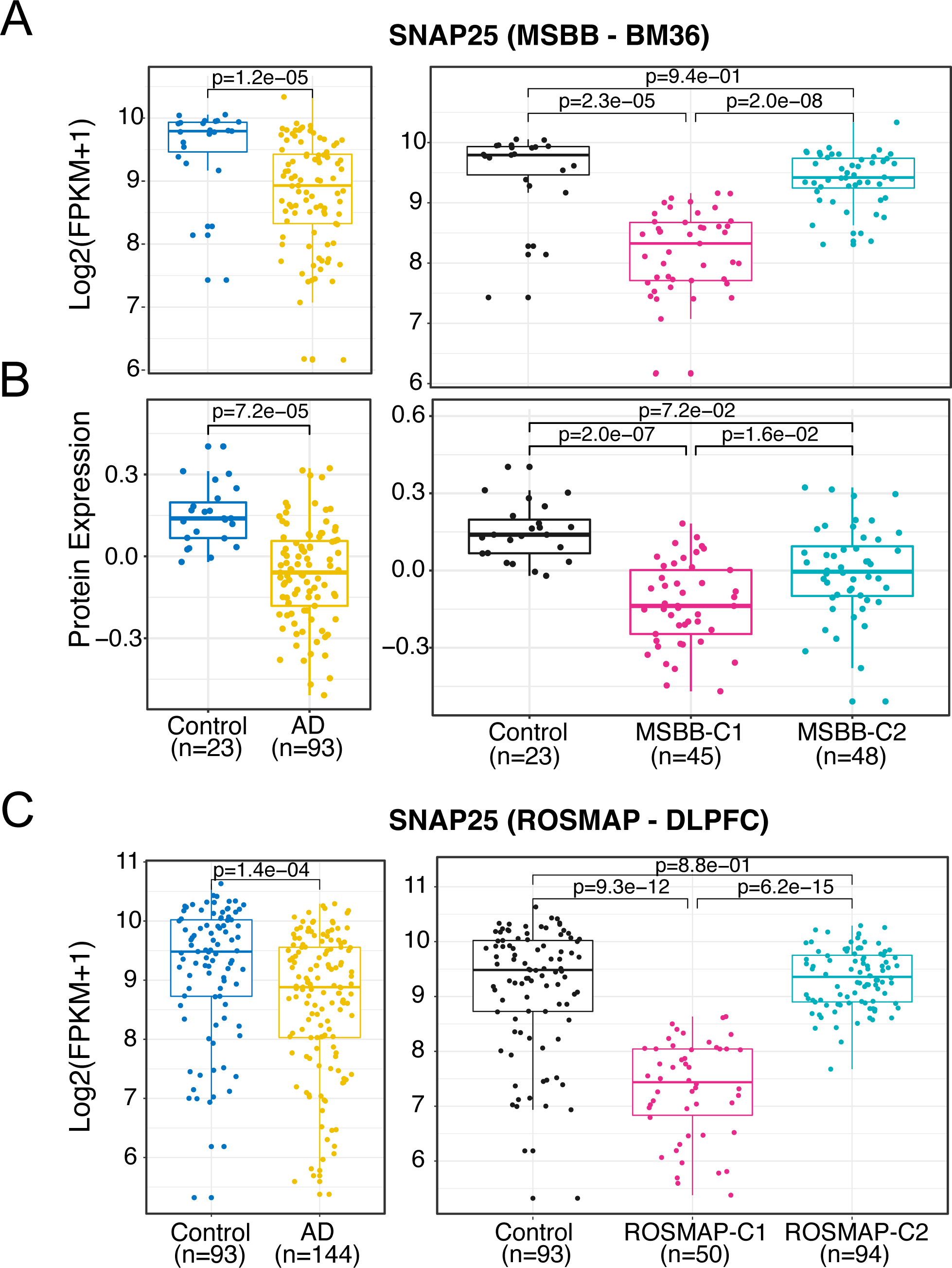
The decreased expression of *SNAP25* in Knight-C4 is replicated in MSBB and ROSMAP cohorts. (**a**) Boxplots showing transcriptomic profiles of *SNAP25* across the two clusters (right) and all ADs (left) in the MSBB (BM36) cohort. (**b**) Boxplots showing proteomic (TMT) profiles of *SNAP25* across the two clusters as mentioned in “a.” (**c**) Boxplots showing transcriptomic profiles of *SNAP25* across the two clusters (right) and all ADs (left) in ROSMAP (DLPFC) cohort.

**S12 Fig.**
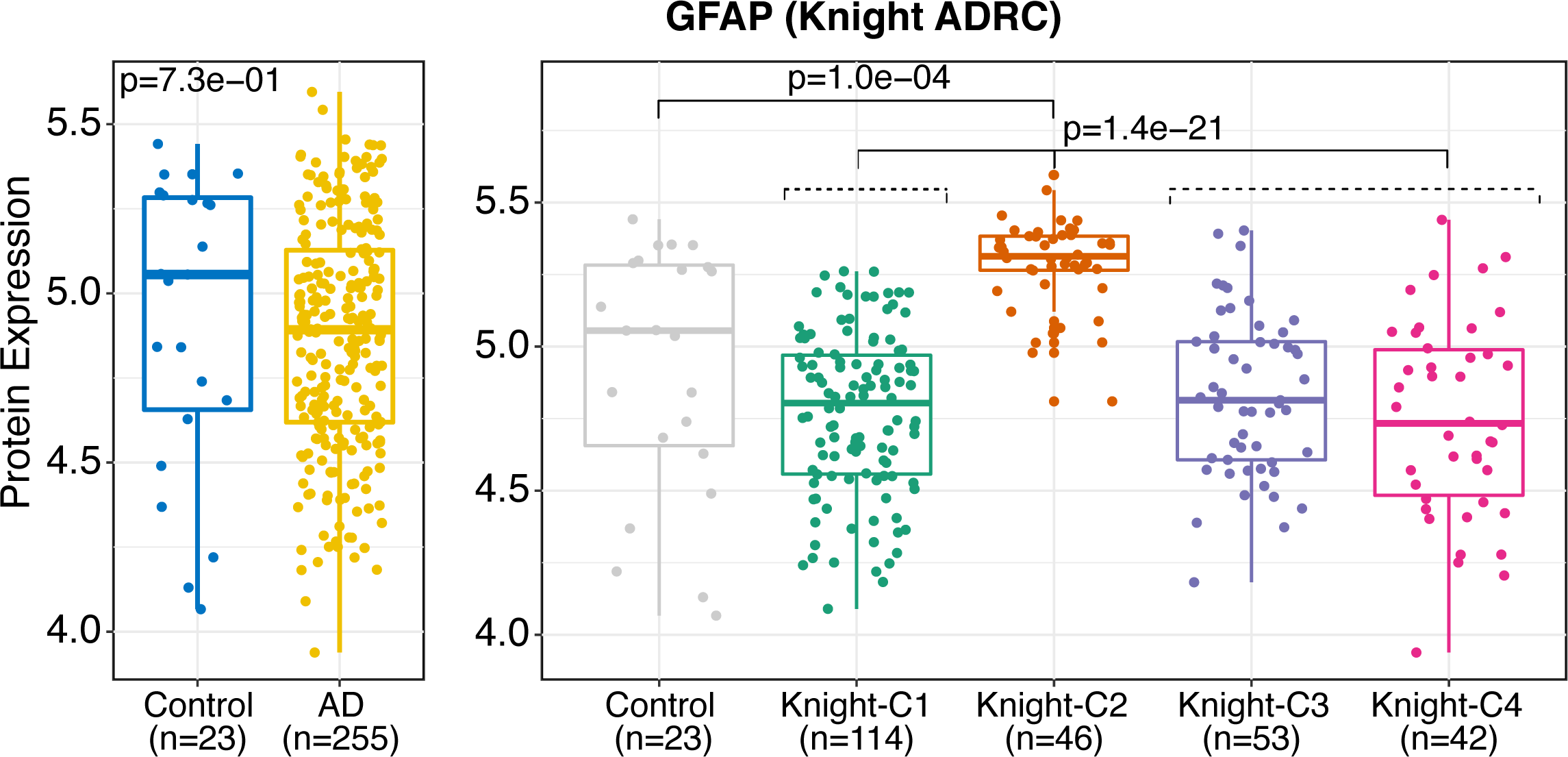
*GFAP* proteomic levels are dysregulated across distinct clusters. *GFAP* protomeric levels are significantly increased in Knight-C2 compared to the control and other AD cases (right).

**S13 Fig.**
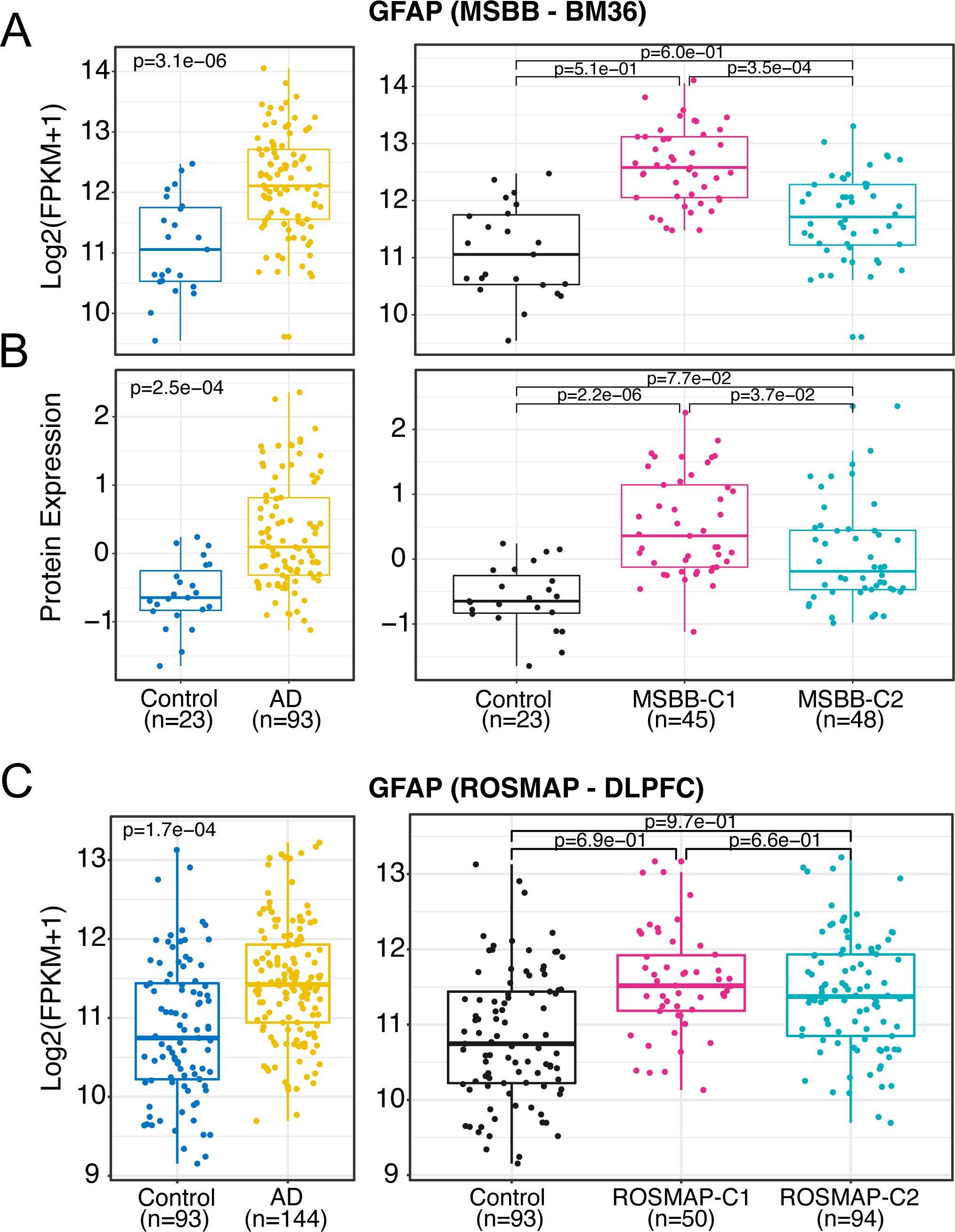
The increased level of *GFAP* is replicated in MSBB and ROSMAP datasets. (**a**) Boxplots showing the transcriptomic profiles of *GFAP* across the two clusters (right) and all AD cases (left) in the MSBB (BM36) cohort. (**b**) Same as “a” but for the proteomic profiles (TMT). (**c**) Same as “a” but in ROSMAP (DLPFC) cohort.

**S14 Fig.**
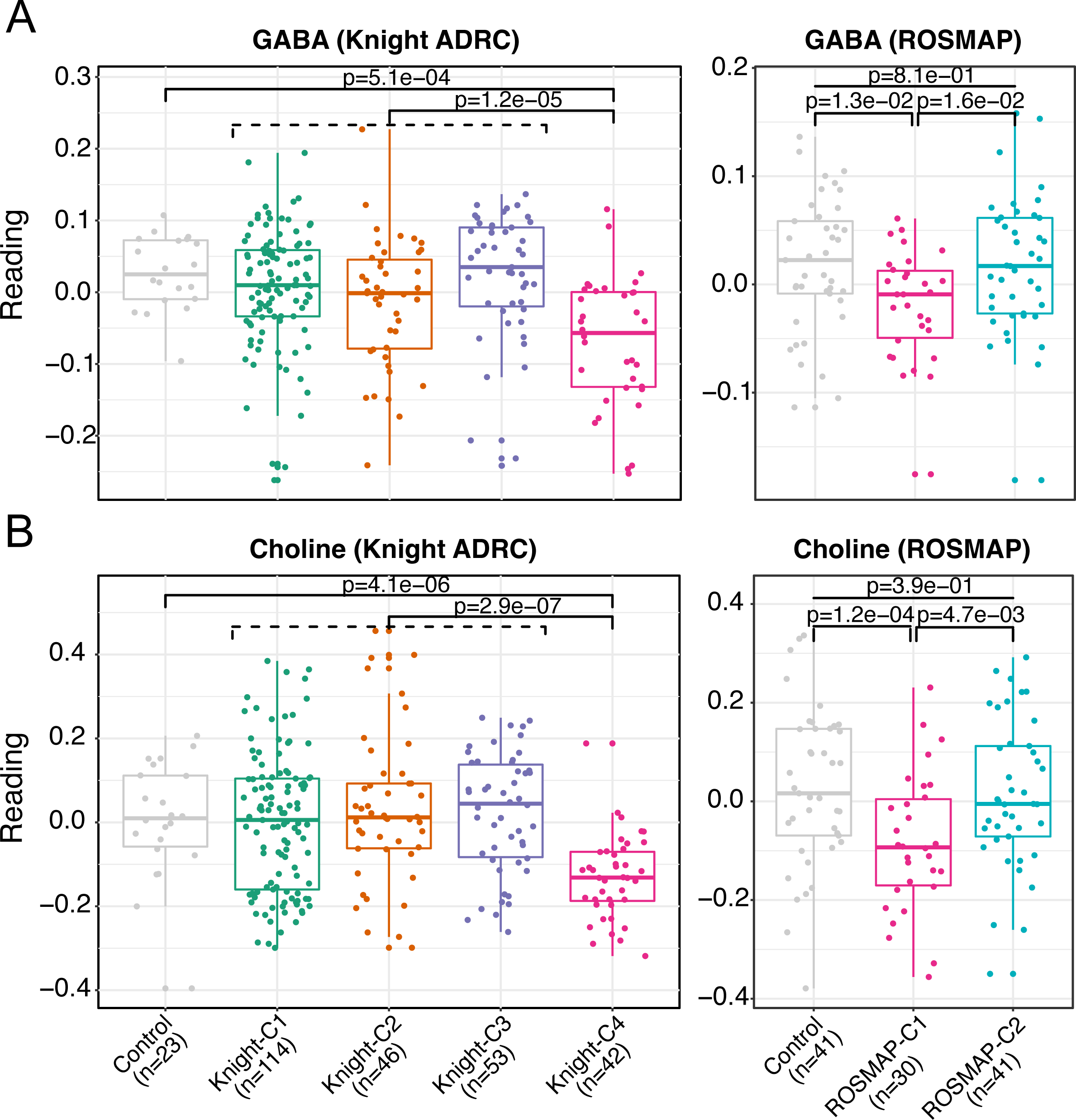
Differential abundance and pathways analyses identified metabolites decreased in Knight-C4 and ROSMAP-C1. (**a**) Boxplots showing the decrease of metabolomics profiles of gamma-aminobutyrate (GABA) in Knight-C4 (left) and ROSMAP-C1 (right). (**b**) Same as “a” but for choline metabolite.

**S15 Fig.**
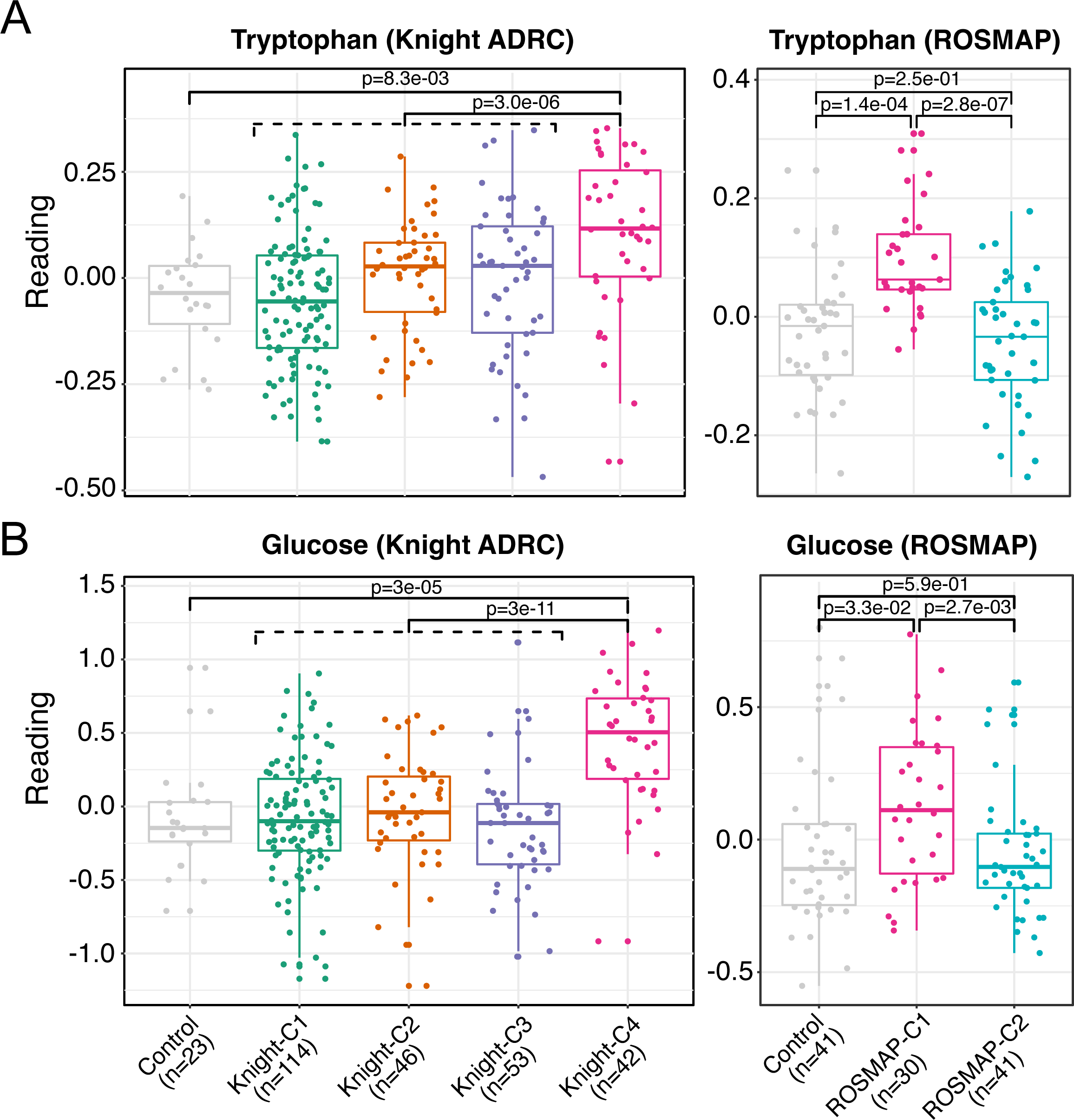
Differential abundance and pathway analyses identified metabolites increased in Knight-C4 and ROSMAP-C1. (**a**) Boxplots showing the increase of metabolomic profiles of tryptophan in Knight-C4 (left) and ROSMAP-C1 (right). (**b**) Same as “a” but for glucose metabolite.

**S16 Fig.**
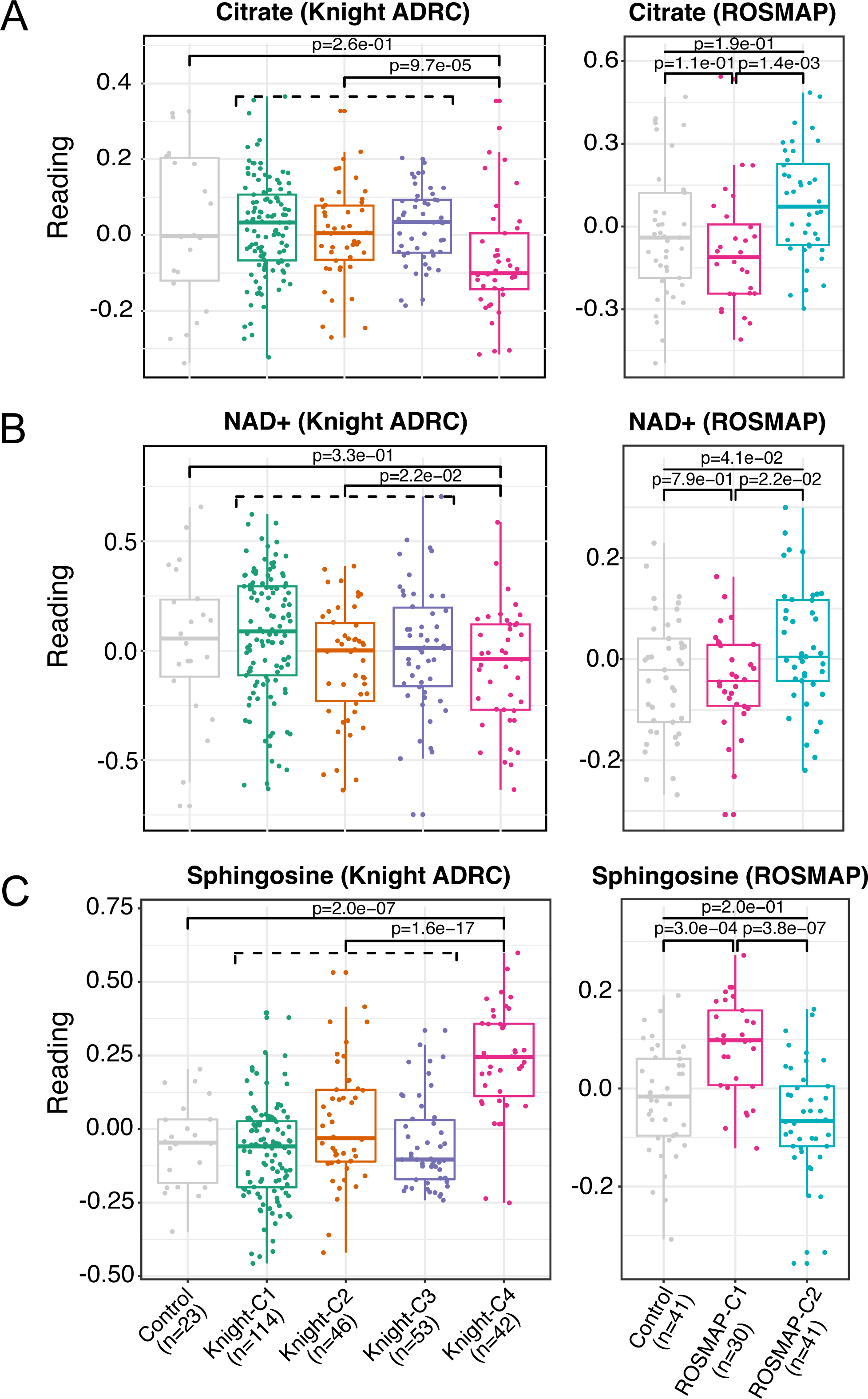
Metabolic differential abundance and pathway analyses identified multiple dysregulated carbohydrate/sugar and sphingolipid metabolites. **(a)** Boxplots showing the decrease of metabolomic profiles of citrate in Knight-C4 (left) and ROSMAP-C1. (**b**) Boxplots showing the decrease of metabolomic profiles of NAD+ in both Knight-C4 (left) and ROSMAP-C1 (right). (**d**) Boxplots showing the increased metabolism levels of sphingosine in both Knight-C4 (left) and ROSMAP-C1 (right).

**S17 Fig.**
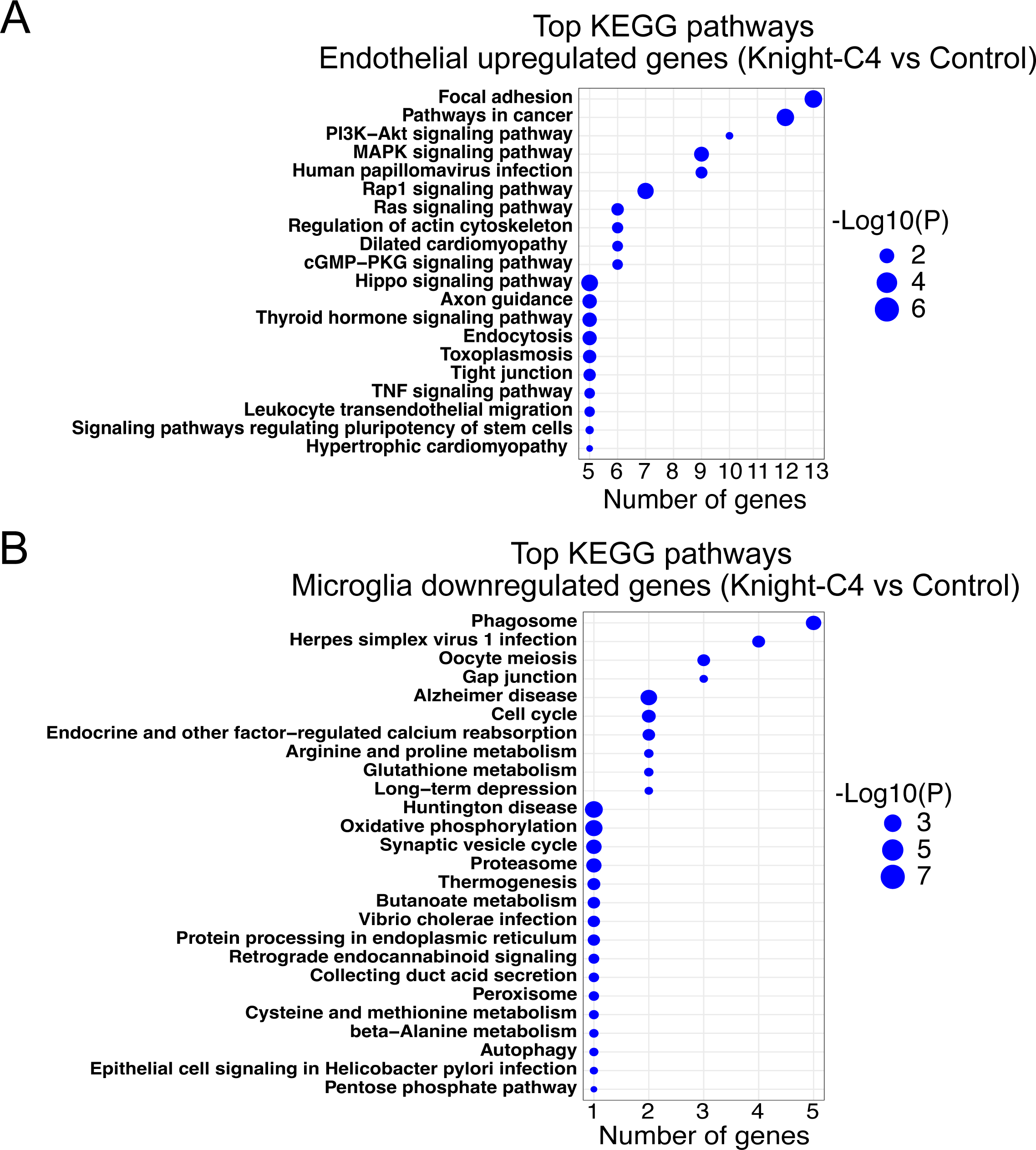
Integrating single-nuclei data with cross-omics profiles identified cell-type specific pathways. **(a)** Top 20 KEGG pathways enriched in endothelial upregulated genes (Knight-C4 vs Control). (**b**) Top 20 KEGG pathways enriched in microglia downregulated genes (Knight-C4 vs Control).

**S18 Fig.**
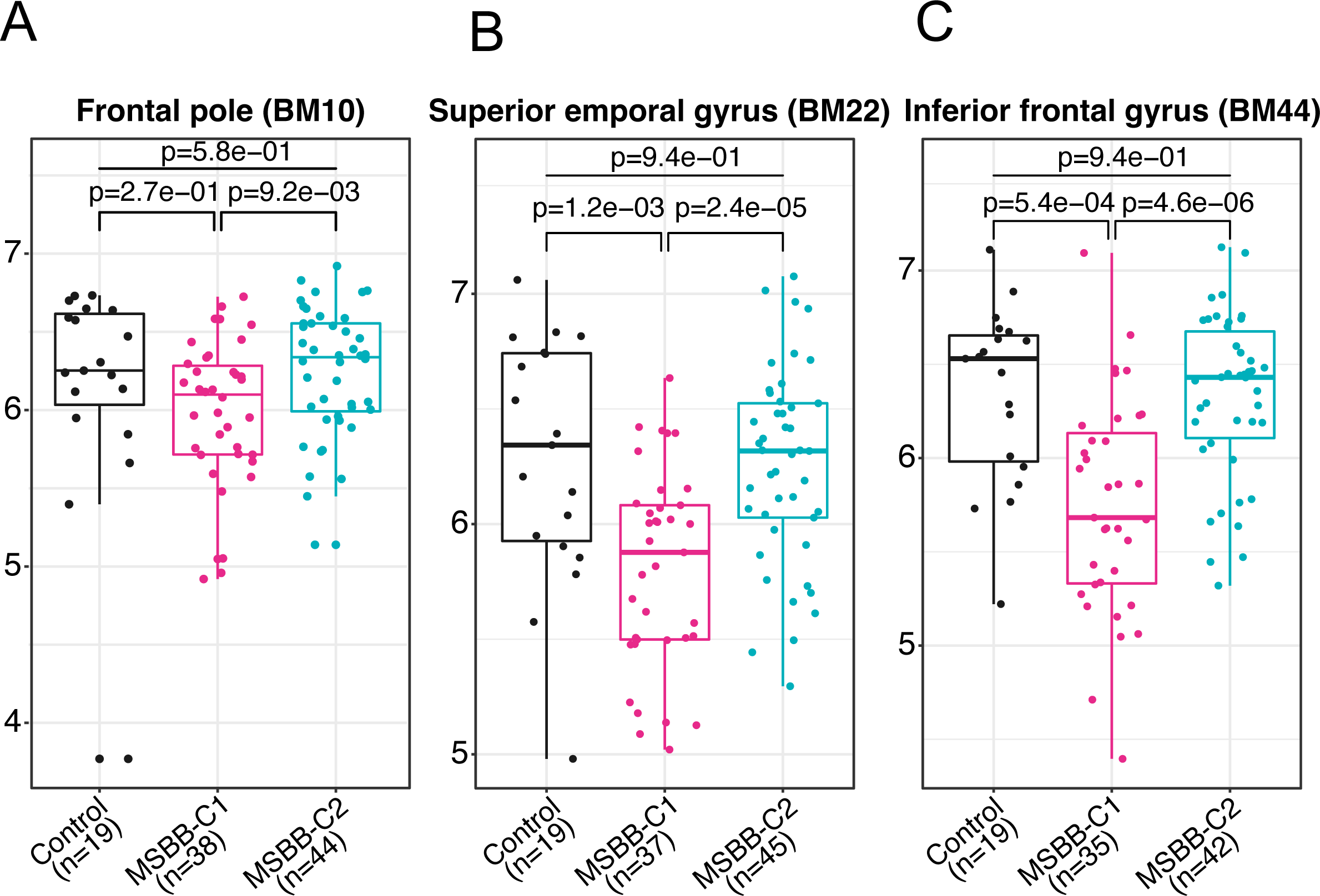
Downregulation of *SNCA* is replicated in additional brain regions from the MSBB study. (**a**) Boxplot showing the transcriptomic profiles of *SNCA* in MSBB-C1 from the frontal pole (BM10) brain region. (**b**) Same as “a” but from superior temporal gyrus (BM22). (**d**) Same as “a” and “b” but from the inferior frontal gyrus (BM44) brain region from MSBB.

**S19 Fig.**
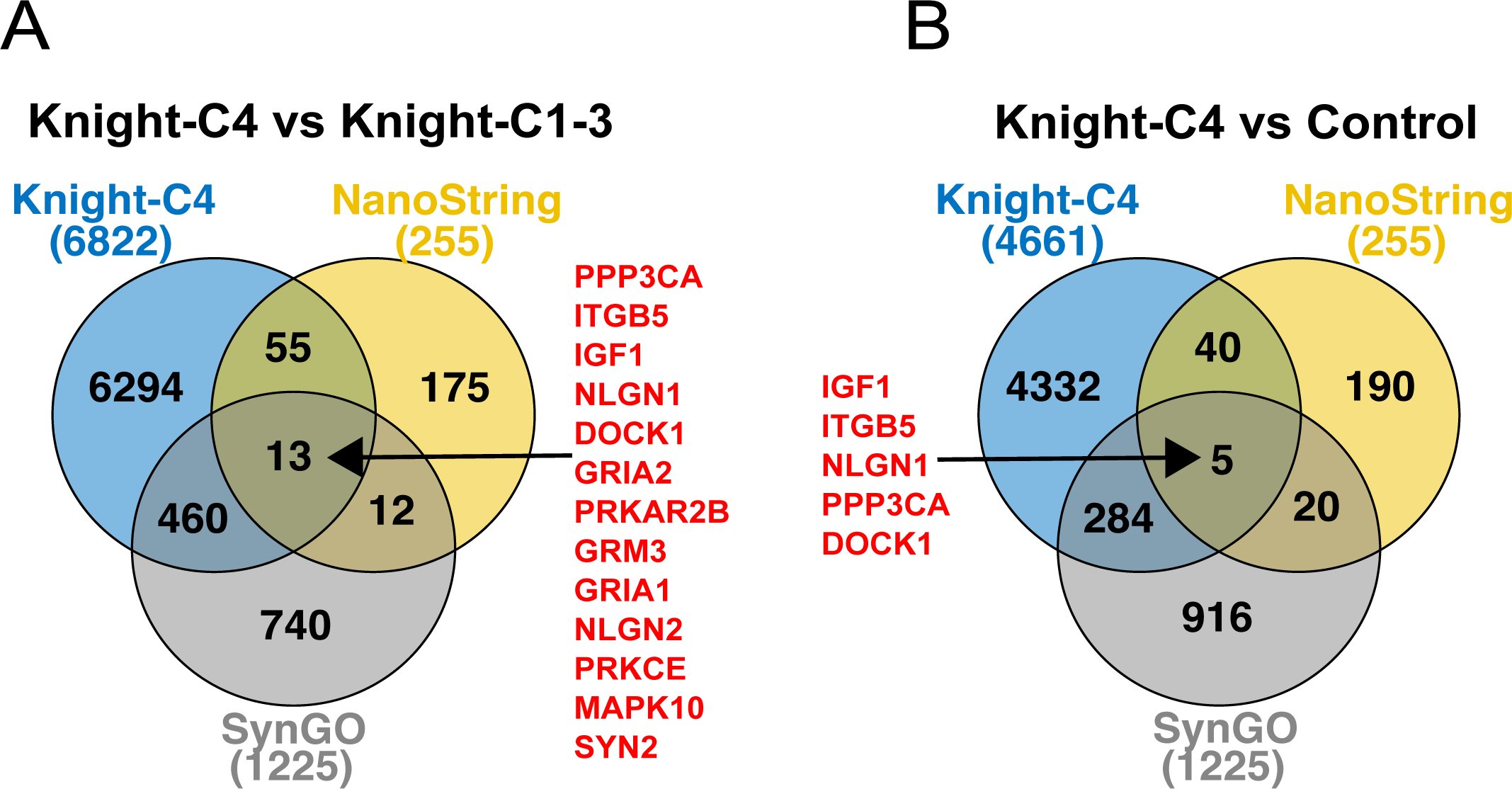
The synaptic dysregulation in Knight-C4 is partially recapitulated in mouse models. **(a)** Venn diagram showing the overlap between significant genes in Knight-C4 compared to other clusters, significant genes from NanoString gene expression data from A53T αSyn mouse models, and synaptic genes from SynGO dataset. (b) Same as “a” but using significant genes in Knight-C4 compared to the control.

**S20 Fig.**
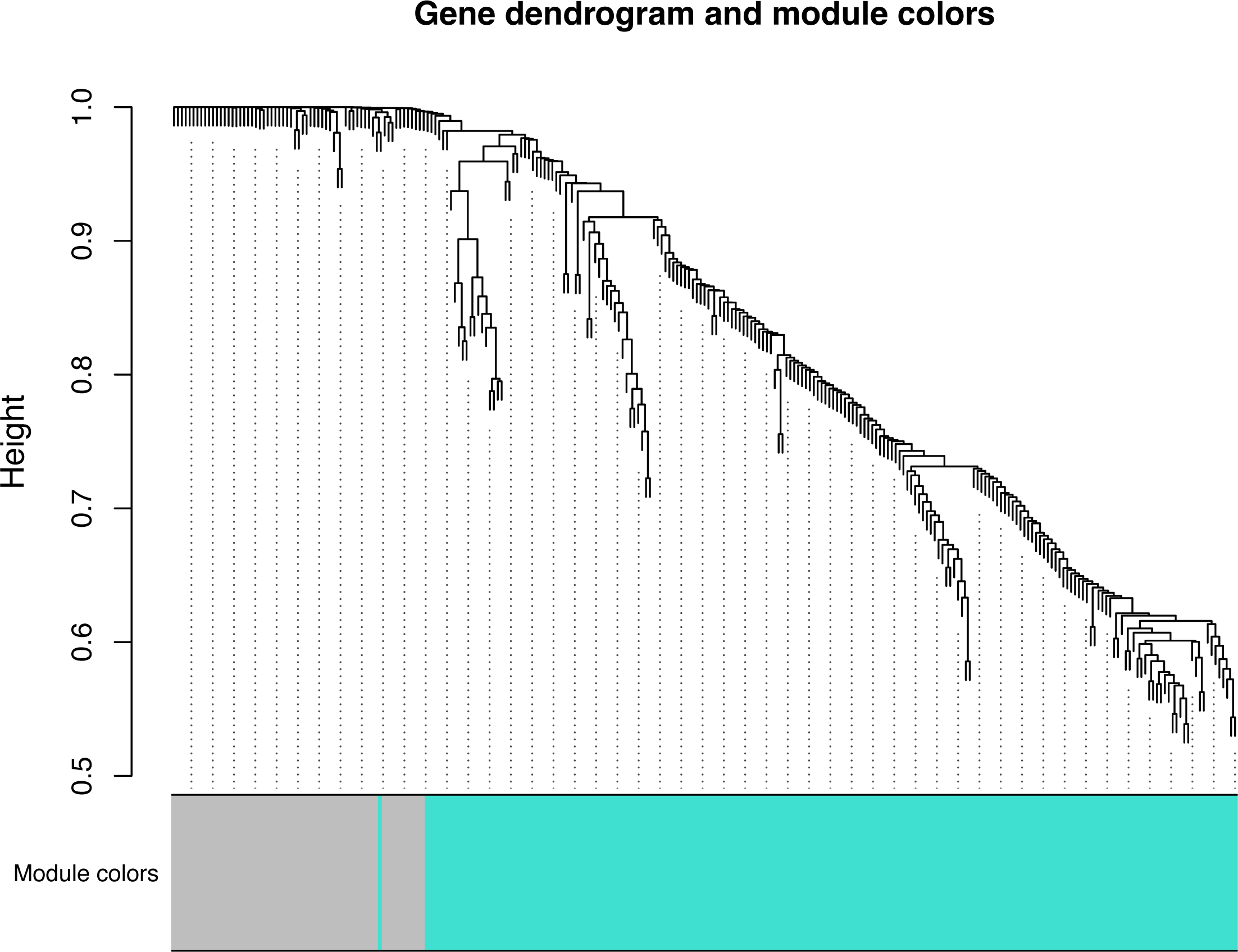
Gene dendrogram and module colors generated by WGCNA showing two distinct gene networks (gray=64, turquoise=209).

**S1 Table.** Demographic characteristics of AD and control cases.

**S2 Table.** Cell proportions for the Knight ADRC, MSBB (BM36), and ROSMAP (DLPFC) cohorts.

**S3 Table.** Differentially expressed genes in the Knight ADRC cohort.

**S4 Table.** Differentially expressed genes/proteins in the MSBB (BM36) cohort.

**S5 Table.** Overlap of significant genes between discovery cohort (Knight ADRC) and replication cohorts (MSBB and ROSMAP cohorts).

**S6 Table.** Differentially abundant genes/metabolites in ROSMAP (DLPFC) cohort.

**S7 Table.** Differentially expressed proteins in the Knight ADRC control.

**S8 Table.** Differentially abundant metabolites in Knight ADRC cohort.

**S9 Table.** Summary of the significant features detected between each cluster and the control.

**S10 Table.** Number of significant features between each cluster and the others.

**S11 Table.** Overlap between significant genes in Knight ADRC, MSBB (BM36), ROSMAP (DLPFC) and synaptic genes.

**S12 Table.** Overlap between differentially abundant metabolites in Knight ADRC and ROSMAP cohorts.

**S13 Table.** Number and percentage of significant features specific to each cell type across four Knight clusters.

**S14 Table.** List of protein-protein interaction (PPI) hub proteins.

